# Myelin pathology in ataxia-telangiectasia is the cell autonomous effect of ATM deficiency in oligodendrocytes

**DOI:** 10.1101/2021.01.22.20245217

**Authors:** Kai-Hei Tse, Aifang Cheng, Sunny Hoi-Sang Yeung, Jia-Nian Ng, Gerald Wai-Yeung Cheng, Qingyang Wang, Beika Zhu, Yong Cui, Liwen Jiang, Julia Kofler, Karl Herrup

## Abstract

Ataxia-telangiectasia (A-T) is a rare genetic disease caused by mutations in the gene encoding the ATM (ataxia-telangiectasia mutated) protein. Although neuronal degeneration in the cerebellum remains the most prominent sign in A-T pathology, neuroimaging studies reveal myelin abnormalities as early comorbidities. We hypothesize that these myelin defects are the direct consequence of ATM deficiencies in the oligodendrocytes (OL) lineage. We examined samples from ten A-T brains in which the ATM mutations had been mapped by targeted genomic sequencing and from *Atm*^-/-^ mice. In healthy human cerebellum, we confirmed the presence of ATM in white matter OLs. In A-T, a significant reduction in OL density was found along with a massive astrogliosis. This white matter pathology was recapitulated in *Atm*^-/-^ mice in an age- and gene dose-dependent fashion. Activated ATM was found expressed both in the nucleus and cytoplasm of OL progenitor cells (OPC) and myelinating mature OL. Its presence in the OL lineage is associated with novel OL-specific functions of the ATM protein affecting all stages of the OL life cycle. Blockage of ATM activity with KU-60019 or inducing DNA damage induced with etoposide altered the cell cycle in self-renewing OPC and triggered ectopic cell cycle re-entry in mature OL *in vitro*. Further, the differentiation program of OPC is highly sensitive to DNA damage either induced directly or by blocking DNA repair. As much of the impact of ATM deficiency in OL is independent of neuronal loss, our findings have important implications for the complex neurological symptoms of human A-T.

**HIGHLIGHTS:**

1. Oligodendrocytes are highly vulnerable to DNA double strand breaks
2. ATM regulates cell cycle control and differentiation of oligodendrocytes
3. Myelin-pathology in Ataxia Telangiectasia is likely the cell-autonomous consequence of ATM deficiency in oligodendrocytes

## Introduction

Ataxia-telangiectasia (A-T) is a rare genetic disease that affects 1: 40,000 to 1:200,000 live births (Salman et al., 2013; Shiloh, 2020; Swift et al., 1986; Taylor et al., 2015). All known cases of A-T are caused by mutations of the gene encoding the ATM protein (Ataxia-telangiectasia mutated), a member of the phosphatidylinositol-3-kinase (PI3K) family that is expressed by all cell types. ATM is best known for its role in coordinating DNA damage response (DDR) to DNA double strand breaks (DSBs) (Savitsky et al., 1995). Once a DSB is formed, a specialized protein complex, MRN (Mre11- Rad50-Nbs1), assembles at the breakage site and serves as a docking site for ATM which then autophosphorylates on serine 1981. The downstream phosphorylation targets of ATM include DNA repair, cell cycle progression and cell death (Paull, 2015). Apart from nuclear functions, ATM serves other physiological functions in the cytoplasm (Boehrs et al., 2007). Indeed, cytoplasmic ATM protein can be found in cytoplasmic vesicles (Barlow et al., 2000; Li et al., 2012; Li et al., 2009; Lim et al., 1998; Watters et al., 1999) and mitochondria (Maryanovich et al., 2012; Valentin-Vega et al., 2012), where ATM participates in a diversity of extranuclear functions independent of genomic repair. These functions include acting as a sensor against oxidative stress (Chen et al., 2003; Chow et al., 2019a; Ditch and Paull, 2012; Guo et al., 2010; Kamsler et al., 2001; Kozlov et al., 2016), as a partner with transcription factors to regulate inflammation (Fang et al., 2014; Hinz et al., 2010; Wu et al., 2006; Wu et al., 2010), and as a regulator of synaptic vesicle trafficking in cortical neurons (Cheng et al., 2021; Cheng et al., 2018) to maintain proteostasis (Lee et al., 2018). The genetic loss of ATM in A-T eventually leads to a regionally variable pattern of neurodegeneration (Borghesani et al., 2000; Kuljis et al., 1997; Shiloh, 2020).

Children with A-T are clinically characterized first and foremost by a progressive cerebellar ataxia (Taylor et al., 2015). The ataxia is linked with cerebellar atrophy that is easily seen on magnetic resonance imaging (MRI), where histopathology confirms a massive loss of cerebellar Purkinje and granule cells (De Leon et al., 1976; Serizawa et al., 1994). Using diffusion tensor imaging, Sahama et al found compromised myelinated tract integrity of corticomotor, corticospinal and somatosensory pathways in adolescent A-T subjects (Sahama et al., 2015; Sahama et al., 2014b). Using MRI-proton spectroscopy, a more recent clinical study also suggested a higher myelin turnover in A-T patients (Dineen et al., 2020). These myelin associated phenotypes in A-T, though less commonly cited, appears in descriptions dating back to some of the earliest neuropathological studies of A-T where demyelination was reported in the posterior column of the spinal cord (Aguilar et al., 1968; De Leon et al., 1976; Sourander et al., 1966; Strich, 1966; Terplan and Krauss, 1969), the cerebellum, optic nerve, corpus callosum (De Leon et al., 1976; Sourander et al., 1966; Terplan and Krauss, 1969) and peripheral nerves (Serizawa et al., 1994). Indeed, white matter (WM) hyperintensities, suggestive of myelin abnormalities, are detectable in A-T patients as early as 17 months of age (Chung et al., 1994; Gouw et al., 2008; McAleese et al., 2017). While the mechanistic connections between ATM deficits and neuronal pathology have been carefully studied in experimental rodent models of A-T (Borghesani et al., 2000; Kuljis et al., 1997), the pathogenesis of myelin pathology in A-T remains largely unknown. In this report, we test the hypothesis that in A-T there is a direct effect of ATM deficiency on the oligodendrocyte (OL) lineage.

Here, we confirm that OL degeneration is associated with ATM deficiency in postmortem A-T brain tissues and *Atm*-knockout mice. Using primary OL culture, we demonstrate that ATM is ubiquitously expressed throughout the OL lineage, and it is required for DSB repair, cell cycle regulation and myelination. Together, we confirm that early myelin defects in A-T are likely primary events of ATM deficiency in OL, rather than a late secondary consequence of neuronal degeneration.

## Materials and Methods

### Materials

All chemicals were purchased from Sigma-Aldrich if not otherwise specified. Cell culture media and supplements were obtained from Life Technologies and Invitrogen of Thermo Fisher Scientific (MA, USA). All pharmacological inhibitors or compounds namely Camptothecin (#1100), etoposide (#1226) and KU-60019 (#4176) were obtained from Tocris Bioscience.

### Postmortem human cerebellar tissues

Formalin fixed paraffin embedded postmortem cerebellar tissues of a normal aging group were kindly provided by Alzheimer’s Disease Research Center (ADRC) Brain Bank at University of Pittsburgh with approvals from the Committee for Oversight of Research and Clinical Training Involving Decedents at University of Pittsburgh. All cases were characterized neuropathologically, and classified as normal by Braak staging, with no known cerebellar diseases. All cases were supplied as tissue sections on microscopic slides at a thickness of 10 μm. Frozen postmortem cerebellar tissues from ten A-T cases, as well as ten age-matched normal controls (NC) were kindly provided by the NeuroBioBank of the National Institutes of Health at The University of Maryland with approvals from Tissue Access Committee at NeuroBioBank. All tissues were isolated from left cerebellar cortex that had been frozen without fixation and stored at -80°C. Tissue samples were embedded in TissueTek O.C.T. medium, cryosectioned at 16 μm, mounted on glass slides and kept at -80 °C until use. Before immunostaining the cryosections were fixed for 30 mins in 4% paraformaldehyde at room temperature. These studies of postmortem tissues were approved by the Committee of Research Practices at The Hong Kong University of Science and Technology (HKUST) as well as Human Subjects Ethics Application Review board at The Hong Kong Polytechnic University (PolyU). The demography of all cases is listed in Table S1.

### Targeted genomic sequencing

To prepare genomic libraries, DNA was extracted using the Qiagen DNeasy Blood & Tissue Kit. After quantifications using Quant-iT™ PicoGreen™ dsDNA Assay Kit on a NanoDrop™ 3300 Fluorospectrometer (Thermo Fisher Scientific), the samples were tagmented, captured, amplified and processed for targeted genomic sequencing using TruSight® Inherited Disease Sequencing Panel (Illumina, United States).. The libraries were normalized, indexed, pooled and quality controlled using High Sensitivity NGS Analysis Kit (DNF-474, Advanced analytical) on Fragment Analyzer^TM^. The pooled libraries were sequenced on an Illumina Mid-Output Kit in NextSeq550 platform in The Biosciences Central Research Facility, HKUST. The raw reads were aligned against the human reference genome (hg19). The aligned reads in the targeted genome were analyzed for single nucleotide polymorphisms (SNPs) or InDels (<u>in</u>sertions or <u>del</u>etions) based on Genome Analysis Toolkit (GATK, Broad Institute). The annotated genetic variants were called and analyzed by the panel-bundled software Illumina VariantStudio^TM^ 3.0. The TruSight® Inherited Disease Sequencing Panel targets 550 genes known to cause rare inherited diseases. A total of ∼30,000 probes spanning 8801 exons and exon-intron boundaries of these genes (Bell et al., 2011), including ATM and other DNA repair genes causing progeria syndromes, were included. The SNPs and their properties such as variant types, genotype (heter/homozygosity), coordinates, exon/intron, protein and cDNA consequences, and potential pathogenicity, were identified. All mutations found in the targeted region were matched against multiple SNP databases including ClinVar (NCBI), COSMIC (Sanger Institute), dbSNPs (NCBI) and HGVS (Human Genome Organization).

### Animal Subjects

Colonies of B6;129S4-*Atm*^tm1Bal^/J (JAX:020943, heterozygous, *Atm*^+/-^; homozygous, *Atm*^-/-^) and C57BL/6J mice (wildtype) were used in this study (Jackson Laboratory, Bar Harbor, Maine). The *Atm*^-/-^ strain carries an engineered mutation in the mouse *Atm* gene which disrupts the multiple exons encoding the kinase domain (Lavin, 2013; Xu et al., 1996). At appropriate age, the animals were deep anesthetized by Avertin (1.25% tribromoethanol, 375 mg/kg, intraperitoneal), the chest cavity was surgically opened and transcardial perfusion with Phosphate buffer saline (PBS) was performed using a peristaltic pump. The animal was then dissected, and its brain was isolated. The left hemisphere was frozen for protein and gene expression analysis, while the right hemisphere was fixed by immersion in paraformaldehyde (4%) for 24 hours at 4 °C. The tissue was then cryopreserved in PBS-sucrose (30% w/v) for 48 hours, followed by embedding and cryosectioning at 10 μm beginning at the midline. All sections were mounted on glass slides which were kept at -80 ^°^C until use.

All animals were housed in a temperature and humidity-controlled environment on a 12 hour light/dark cycle with food and water *ad libitum*. All animals were maintained and cared for by the Animal and Plant Care Facility (APCF) at HKUST in compliance with Legislations and The Code of Practice for Care and Use of Animals for Experimental Purposes of Hong Kong. All animal experiments and analysis were approved by both the Animal Ethics Committee, of the Committee on Research Practices of HKUST and Animal Subjects Ethics Sub-Committee of PolyU. All procedures were also conducted with a license from the Department of Health, Government of Hong Kong.

### Primary oligodendrocyte culture

Mice at postnatal day 2 (P2) to P6 mice were sacrificed by decapitation and brains dissected free of the skull (Emery and Dugas, 2013; Luo et al., 2016; Tse et al., 2018). The cerebellum and olfactory bulbs were removed while whole cerebrum was minced with fine scissors, followed by trypsin-based enzymatic digestion (0.25%, 30 mins, 37°C). After neutralization, the cell suspension was filtered through a cell strainer with a 40 μm pore size. The suspension was then centrifuged (1500 rpm, 5 min), the cell pellet was resuspended, and the cells transferred to a dish coated with BSL1 (1:500, L1100, Vector Laboratories) for 15 min at room temperature to remove any isolectin B4^+^ microglia. The resulting suspension, consisting primarily of a mixture of OL and astrocytes, was collected, centrifuged and plated on poly-L-lysine coated culture dishes to expand the OL progenitor cell (OPC) population. OPC growth medium was composed of DMEM/F12 (Gibco), fetal bovine serum (FBS, 1% v/v) and N2 supplement (bovine serum albumin [70 μg/mL], insulin [5 μg/mL], human transferrin [5 μg/mL], putrescine [1.6 μg/mL], progesterone [60 ng/mL], sodium selenite [5 ng/mL] and L-thyroxine [400 ng/mL] – Sigma). Platelet derived growth factor-AA (PDGF, 10 ng/mL, Sigma), NT3 (1 ng/mL, Peprotech) and CNTF (10 ng/mL, Peprotech) were added to stimulate OPC proliferation. To differentiate OPC into mature OLs (mOLs), the expanded OPCs were harvested after 5 days in vitro and plated on coated 12 mm glass coverslips (35,000 cells each). The OPC were then induced to differentiate for 7-14 days using 34 ng/mL triiodothyronine (T3) in the absence of growth factors, and at reduced FBS (0.1%) concentration. The DNA synthesis at the S phase was detected by Click-iT™ EdU Cell Proliferation Kit for Imaging according to manufacturer’s instruction (ThermoFisher)

### Oli-Neu cell culture and transfection

An OPC cell line, Oli-Neu, was kindly provided by Dr Jacqueline Trotter (University of Mainz, Germany). Oli-Neu cells retain an OPC phenotype but can differentiate into myelin-expressing OL upon db-cAMP stimulation (1 mM) (Jung et al., 1995; Pereira et al., 2011; Sohl et al., 2013). Oli-Neu cells were cultured in DMEM/F12 medium supplemented with 1% normal horse serum, 1% N2 supplement and 1% penicillin-streptomycin. For shRNA experiments, cells were plated on coated 13 mm glass coverslips (20,000-35,000 cells) in a 24 well plate. To introduce shRNA, cells were then transfected with a total of 0.5 – 1 µg of DNA construct (GFP-*Atm*-shRNA (TL320267) or GFP-*Atr*- shRNA (TL519184) or a scrambled 29-mer control shRNA cassette in pGFP-C-shLenti vector (OriGene) using Lipofectamine 2000 reagents (Thermo Fisher Scientific). The transfected cells were examined under a fluorescent microscope 48 hours after transfection before proceeding with immunocytochemistry or gene expression experiments.

### Primary neuronal cell culture

Primary cortical neurons were derived from wildtype mouse embryos, as reported earlier (Cheng et al., 2018; Cicero and Herrup, 2005). Briefly, E16 embryos were isolated, and the cerebral cortices were dissected after the meninges were removed. The cortices were then enzymatically dissociated in trypsin-EDTA (1X, 0.25%, Gibco) for 12 min at 37 ^°^C. After neutralization with 10% fetal bovine serum in DMEM, the cells were transferred to NeuroBasal medium containing B27 supplement (2%), GlutaMAX (1%) and penicillin-streptomycin (1%, 10,000 U/mL; all from Life Technologies). The tissue suspension was triturated through a 10 mL glass pipette 10 times before being allowed to settle by gravity in a 15 mL conical tube for 8 minutes. The supernatant containing dissociated cells was transferred to another conical tube for quantification. Neuronal cells were plated at a density of 8,500 cells/cm^2^ on poly-L-lysine coated glass coverslips in 24 well plates for microscopy experiments. All cultures were kept at 37 °C in a humidified incubator with 5% CO2/95% air. Half of the medium was removed and replaced with fresh medium every two days up to 14 DIV (days *in vitro*).

### Immunohistochemistry and immunocytochemistry

For immunohistochemistry of the human paraffin sections, slides were baked for 1 hour at 60 ^°^C before being deparaffinized twice in xylene for 10 min. After serial rehydration in 100, 95, 70 and 50% ethanol, epitope retrieval was performed at 100 ^°^C water bath in basic Tris-EDTA buffer (10 mM Tris Base, 1 mM EDTA solution, 0.05% Tween 20, pH 9.0) for 30 min. Any residual peroxidase activity was quenched by incubation in 3% H2O2 for 10 min. The slides were blocked with normal goat or horse serum for 30 min before application of primary antibodies (anti-Olig2, 1:300, anti-ATM(2c1), 1:300) overnight at 4 ^°^C. All primary antibodies used for immunohistochemistry are listed in Table S2. After TBS washes, sections were incubated with the corresponding secondary antibody (VectaStain® Elite® ABC HRP Kit, Vector laboratories) according to the manufacturer’s protocol. Antibody binding was visualized by 3,3’-diaminobenzidine (DAB Peroxidase HRP Substrate Kit, Vector laboratories) and all tissue was counter stained with Meyer’s hematoxylin. All tissue sections coverslipped with DPX mounting compound before viewing.

For immunofluorescence, after PBS washes, the tissues were blocked with normal donkey serum (10%) in PBS containing Triton X-100 (0.3%) for 1 h at room temperature. Then tissues were incubated with specific primary antibodies overnight at 4 ^°^C. After further PBS washes, the specimens were incubated with secondary antibodies conjugated with Alexa Fluor 488, 555 or 647 fluorochromes (Life Technologies) for 1 h at room temperature, and counterstained with DAPI (4’,6-diamidino-2- phenylindole). The sections were then coverslipped with Hydromount (National Diagnostics). All tissue sections were examined and imaged on an upright microscope (BX53 with DP80 camera, Olympus) and imaged with 20x (UPlanSApo, 0.75 N.A.) or 40x objectives (UPlanSApo, 0.95 N.A., all Olympus) using an X-Cite® 120Q fluorescence illuminator (Excelitas Tech Corp) and appropriate filters.

For immunocytochemistry, OLs were cultured on coverslips in 24-well tissue culture plates and fixed with paraformaldehyde (4%) for 20 min after washing with PBS. The fixed cells were permeabilized using 0.3% Triton X-100 and blocked with normal donkey serum (5%) in PBS for 30 min. Primary antibodies were applied for 2 h at room temperature followed by PBS washes and fluorochrome conjugated secondary antibodies. Negative controls were collected by omitting the primary antibody. For high resolution imaging of the nucleus, specimens were visualized on a Leica TCS SP8 confocal laser scanning platform equipped with Leica HyD hybrid detector and visualized through a 63x/1.40 N.A Oil lens (Leica HC PL APO CS2). In each experiment, 0.3-0.5 μm optical sections were imaged with constant laser power. The Z-projection of the entire optical stack was presented as a single image.

### Transmission electron microscopy

The brain regions of interests (cortex, corpus callosum and cerebellum) were isolated from *Atm*^-/-^ mice and their wildtype littermates for fixation, processing, ultrathin sectioning, and TEM imaging as described earlier (Cui et al., 2014). Brain samples were diced into small pieces before immediately being frozen in a high-pressure freezing machine (EM HPM100; Leica). Specimens were ultrathin sectioned at 70 nm thickness using a Leica UC7 ultramicrotome. The sections were mounted on copper grids then stained/contrasted with aqueous uranyl acetate-lead citrate and visualized by Hitachi H7650 transmission electron microscope (Hitachi High-Technologies).

### Image analysis

For human tissue, six randomized images (690 x 520 μm) were taken from cerebellar gray matter (GM, spanning the molecular, Purkinje and granule cell layer) and cerebellar white matter (WM, folia and deep regions). The total number of immunoreactive cells were counted per image, and their density calculated. For mouse tissues, three images (690 x 520 μm) covering the neocortex (Cx), corpus callosum (CC) and cerebellum (CBM) were taken from three serial sagittal sections for analysis. The average count from the triplicates was taken as the density of each marker per region. For cultured cells, at least than 100 nuclei per field were counted for each condition.

### Western blotting

The left neocortex of each mouse or a human tissue section removed from a slide was lysed for protein extraction using RIPA buffer (radioimmunoprecipitation assay buffer) supplemented with phosphatase and protease inhibitors (Roche Diagnostic). Protein sample concentrations were quantified by bicinchoninic acid assay (Bio-Rad) and normalized amounts of proteins (20 - 50 μg) were electrophoresed on a 10 - 15% SDS-polyacrylamide gels. After transfer to PVDF membranes, non-specific binding was blocked with 5% non-fat milk followed by primary antibody incubation (Table S2). The membranes were probed with horseradish peroxidase-linked secondary immunoglobulins (Cell signalling) before visualization with chemiluminescent substrates (SuperSignal™ West Pico, Dura or West Femto Substrates, Thermo Scientific). The chemiluminescent signals were detected by blue sensitive medical X-ray film, and the resulting band densities were digitalized for quantification using ImageJ software. GAPDH was used as the protein loading control.

### Gene expression

The gene expression analysis of post-mortem human tissue, mouse tissue and cultured cells was performed as described (Tse et al., 2018). Total RNA from frozen human cerebellar sections, dissected mouse brain regions or four wells of cultured cells were extracted using RNeasy mini kit (Qiagen). All contaminating genomic DNA was cleared by DNase I (1 U/μL). All cDNA samples were reversed transcribed using High-capacity RNA-to-cDNA kit (Applied Biosystem). Gene expression analysis was performed using Fast SYBR® Green Master Mix (Applied Biosystem) in a Light Cycler 480 (Roche) using the human or mouse specific primers optimized by PrimerBank (Spandidos et al., 2010). All gene expression levels were calculated relative to housekeeping genes (18S, β-actin and GAPDH). Data analysis was performed using RT² Profiler PCR Array Data Analysis (Qiagen) based on the 2^-ΔΔCt^ method (Livak and Schmittgen, 2001).

### Statistical Analysis

At least three independent experiments were performed, and all data are presented as mean value ± SEM. For pairwise comparisons, unpaired t-tests were performed. For comparisons between multiple groups with one variable, one-way ANOVA with Dunnett’s or Tukey’s multiple comparisons test was performed. For comparisons between multiple groups with two variables, a two-way ANOVA with Holm-Šídák’s multiple comparisons test was performed. The correlations between observations were tested by Pearson correlation coefficient test. All statistical analyses were performed using GraphPad Prism software version 10.00 (GraphPad Software Inc.). The statistical significance level was set as P < 0.05.

## RESULTS

### The loss of oligodendrocytes in human A-T cerebellum

We first confirmed the pathology of neurons and OL lineage in post-mortem cerebellar tissues from ten A-T subjects, where the pathogenic ATM mutations were confirmed by targeted genomic sequencing (Table S1). In the cerebellum, a classic atrophy of molecular, granule cell and Purkinje cell (PC) layers. We found that PC soma size was significantly reduced in each A-T case. The overall PC density, however, only trended lower with considerable variability (Fig.1A, B). We plotted both density and soma size variables as a function of age. This analysis reveals that there was a significant loss of PC density with age in the A-T subjects, but not in control cases. The reverse was true for PC soma size which decreased with age in normal controls with little changes in A-T samples (Fig.1C).

**Figure 1.**
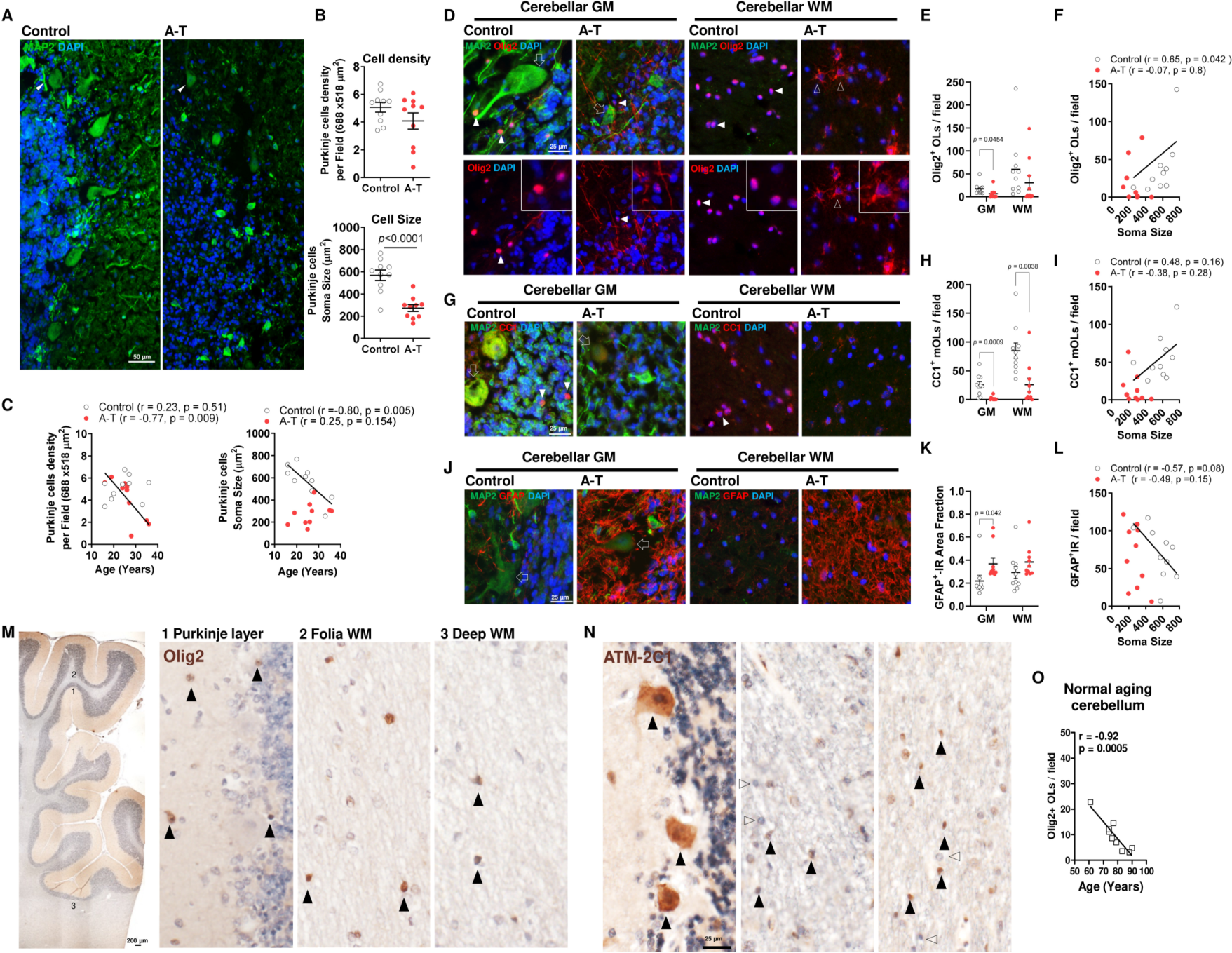
Loss of cerebellar neurons and OLs in Ataxia Telangiectasia **A** Immunohistochemistry of MAP2 showing the cerebellar neurons in Control (n = 10) and A-T subjects (n = 10). A-T cerebella showed classic atrophy with reduced number, reduced soma size and dysplasia of Purkinje cells (PC, Green, MAP2, arrowhead). **B** Quantifications of PC density and soma size with a significantly reduced soma size Purkinje cells in A-T subjects (unpaired t-tests, p-value on graph). **C** Plots showing the significant negative correlation between PC density and age in A-T and the significant negative correlation between Purkinje soma size and age in control subjects. **D** Representative image of Olig2-MAP2 immunohistochemistry showing PC degeneration (open arrows) and the Olig2+ OL population (filled arrowheads) in the GM (PC layer, left) and WM (deep nuclei and arbor vitae, right) of control and A-T cerebellum (upper panel). Olig2-immunoreactivity (IR) resembling fibrillary astrocytes were identified in cerebellar GM and WM of A-T but not controls (Open arrowhead, see inset for higher magnifications). **E** Quantification showing the significant reduction of Olig2+ OL in cerebellar GM (unpaired t-tests, p-value on graph). **F** Plot showing a significant positive correlation between Olig2+ OL density and PC soma size in control but not in A-T. **G** Representative image of CC1-MAP2 immunohistochemistry showing PC degeneration (open arrows) and the CC1+ mOL population (filled arrowheads) in the GM and WM of control and A-T, as in **D**. **H** Quantification of CC1+ mOL in cerebellar GM and WM. **I** Plot showing the significant positive correlation between CC1+ mOL density and PC soma size in control but not in A-T. **J** Massive astrogliosis was identified throughout A-T cerebella with excessive GFAP-IR across layers **K** Quantifications of GFAP+ IR area showed significant astrogliosis in A-T but not control cerebellum. **L** Plot showing the significant negative correlation between GFAP-astrogliosis and PC soma size in control, but not in A-T. **M, N** To confirm OL express ATM in normal cerebellum, Olig2+ OL was traced in a normal aging cohort cerebellum. Olig2+ OL was found throughout cerebellar layers (brown, filled arrowheads, PC layer, folia WM and deep WM), and ATM-IR (antibody 2C1, filled arrowheads, brown; negative glial cells, open arrowheads) was found in the nucleus and cytoplasm of Purkinje cells as well as in nucleus resembling Olig2+ OL throughout cerebella.

In addition to these changes in the neurons of the A-T cerebellar cortex, we also observed a remarkable reduction of the OL population. We marked cells at all stage of OL maturation by immunostaining for Olig2, a pan-OL marker and OL-specifying transcription factor (Fig. 1D). The loss of nuclear Olig2^+^ cells was significant in the cerebellar gray matter (GM) but did not reach statistical significance in the white matter (WM, Fig. 1E). The myelinating and mature OL (mOL) population was also significantly reduced in the A-T cerebella. Using CC1 as a marker [anti-APC, clone CC1, (Bin et al., 2016), Fig. 1F], we identified a 92.1% and 69.6% reduction of mOL in the GM and WM, respectively (Fig. 1G). More dramatic than the reduction in Olig2^+^ cell number, there was a noticeable shift in the cellular location of Olig2 from nucleus to cytoplasm in the A-T cases. Indeed, the morphology of the Olig2 cells more closely resembled that of astrocytes than normal OL cells (Fig. 1D, lower panel). In keeping with this observation, GFAP (glial fibrillary acidic protein) immunohistochemistry revealed a major astrogliosis in the A-T cerebella both in the GM and WM (P = 0.0177, Fig. 1H, I). These observations suggested that the OL progenitor cells (OPCs) were adopting an astrocytic cell fate by shuttling Olig2 to the cytoplasm, as reported by others (Cassiani-Ingoni et al., 2006; Zhao et al., 2009; Zhu et al., 2012; Zuo et al., 2018). Taken together, the data suggest that OL lineage is disrupted in A-T cerebellum.

To determine whether OL degeneration in A-T cerebellum was a direct effect on OLS or an indirect effect secondary to the well documented neuronal changes, we correlated the neuronal and OL histopathology data (Fig1. J-L). A significant positive correlation between Olig2^+^ OLs and Purkinje cell size was found in normal control but not A-T. Although no correlation between astrocytes, CC1^+^ mOL and neurons was found in any groups, the opposing trends of Olig2^+^ OLs suggested that the overall OL degeneration in A-T may not be a secondary consequence of neuronal atrophy or death.

ATM expression in human cerebellum has been previously described in PC and its function there is well documented (Li et al., 2011; Shiloh, 2020). Yet, few, if any, studies have described the ATM expression in the glial cells in the human brain. Using Olig2, we identified the OL population in FFPE cerebella tissues from an aging cohort diseased from known neurological disease (Fig. 1M). We found that the cerebellar Olig2^+^ OL population was distributed across the PC layer, cortical WM and deep WM in the cerebellum, their density decreased gradually during normal aging (Fig. 1N). Using a well-characterized ATM antibody (clone 2C1), we confirmed the nuclear and cytoplasmic ATM expression in PC (Li et al., 2011), and identified a strong nuclear ATM expression in a significant number of small and dark nuclei along the myelin fiber tracts (Fig. 1O). These ATM expressing cells throughout the cerebellar WM were highly reminiscent of the pattern of Olig2^+^ OL cells. This putative ATM expression in OL population was confirmed in three independent single cell RNA-seq databases of human and mouse cerebellum and brain tissues (Aldinger et al., 2021; Kozareva et al., 2021; Sjostedt et al., 2020), as in Fig. S1.

### Myelin deficits in young Atm-knockout mice

Having confirmed ATM expression in OL and its demise in human A-T, we turned to *Atm*- knockout mice. Blackgold II myelin staining showed that intracortical myelin fibers in the frontal cortex (Cx) and myelin tracts in the corpus callosum (CC) were reduced in *Atm*^-/-^ animals as early as 1 month of age (MO). Curiously, in *Atm^-/-^* cerebellum (CBM) we detected no changes in Blackgold II staining. (Fig. 2A). The four major myelin proteins (MAG, MBP, PLP and MOG) were observed with immunohistochemistry. No remarkable myelin fiber pattern changes were observed in the CC. However, a reduction of MAG, MBP and MBP were seen in the CX and CBM of *Atm*^-/-^ mice (Fig.4B). To quantify these reductions, myelin protein levels in the CX and CBM were measured by immunoblotting. Indeed, *Atm-*knockout genotype contributed to a significant reduction of MAG (P < 0.0012), MBP (P < 0.0268) and PLP (P < 0.0119) across brain tissues (Fig. 2C, D). In particular, MAG and MBP were significantly reduced in the CX and CBM of *Atm*^-/-^ mice, respectively (Fig. 2E). In the myelin structure (Fig. 2F), MAG is the glycoprotein that ties myelin ensheathment to axons in the periaxonal region, while MOG is the glycoprotein on the OL cytoplasm. MBP, together with PLP, forms the lipid-rich dense bands of the compact myelin. The loss of MAG and MBP suggested that the integrity of the myelination is compromised in *Atm^-/-^*.

**Figure 2.**
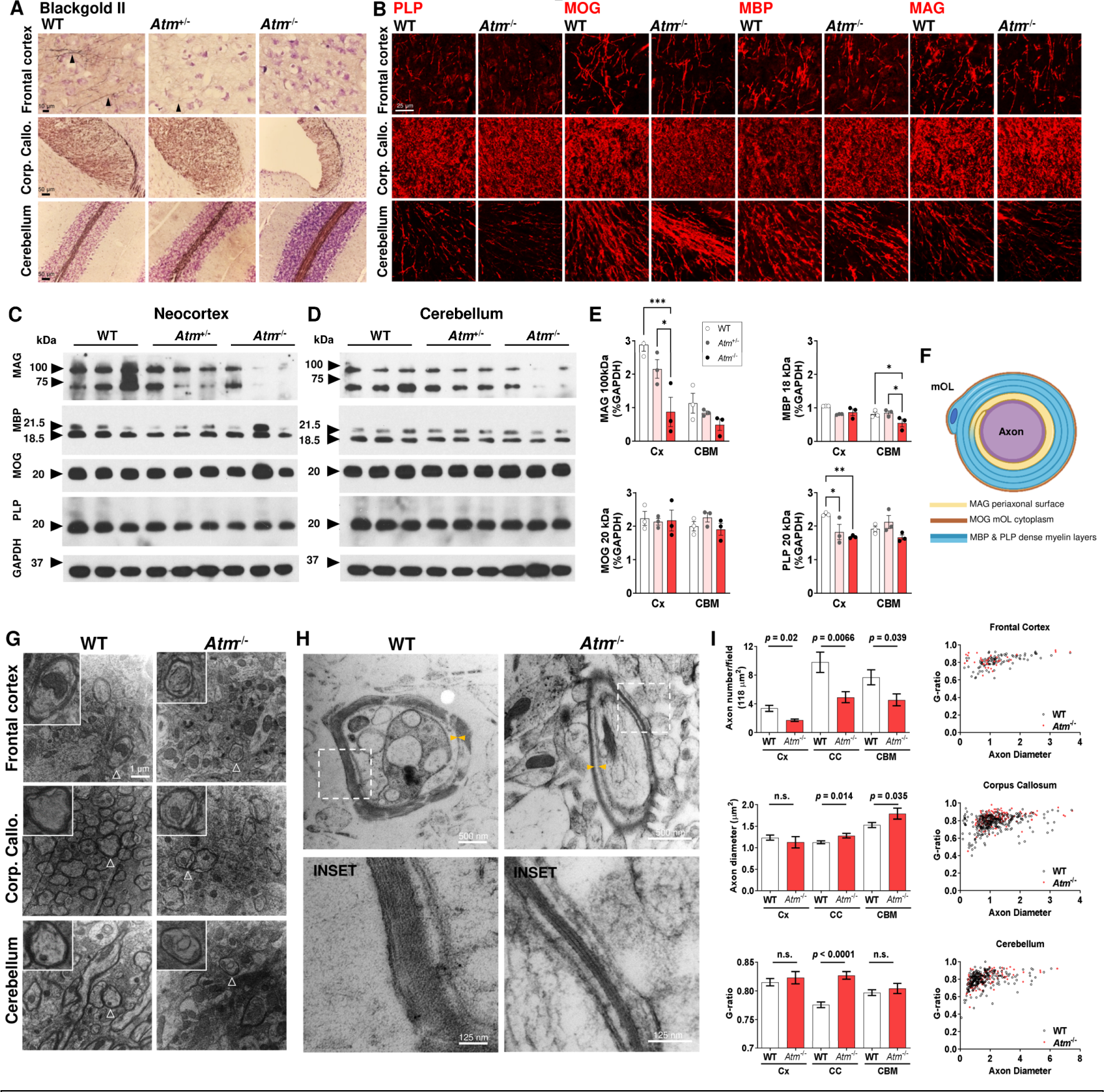
Myelin abnormalities across brain regions in *Atm*-/- mice **A** Representative Blackgold II myelin stain showing a significantly reduced intracortical myelin fibers in the frontal cortex of *Atm*+/- and *Atm*-/- mice at 1 month of age. The size of corpus callosum was also reduced *Atm*-/- mice, while the changes of cerebellum were minimal in this special stain. **B** Representative immunohistochemistry images of myelin proteins (MAG, MBP, MOG and PLP) in the cortex, corpus callosum and cerebellum of wildtype and *Atm*-/- at 1 month of age. **C, D** Immunoblots of myelin proteins (MAG, MBP, MOG and PLP) in the neocortex and cerebellum showing *Atm*-/- genotype is associated with a major change of MAG (p = 0.0012), MBP (p = 0.0268) and PLP (p = 0.0119), with quantification against GAPDH as shown in **E** (Two-way ANOVA with Holm-Šídák’s multiple comparisons test; *p < 0.05, **p < 0.01, ***p < 0.001). **F** The normal distribution of MAG, MBP, MOG and PLP in the myelin sheath is indicated. **G** Transmission electron microscopy confirmed myelin abnormality in the neocortex, corpus callosum and cerebellum of the *Atm*-/- mice, with representative myelinated axon magnified in insets. **H** At high power, axon with dysmyelination or poorly compacted myelin layer was observed in *Atm*-/- mice (right). **I** TEM image quantification showed a significant reduction of axon density (top), diameter (middle) and g-ratio (bottom) across brain regions (unpaired t-test, p-value on graph, 10-14 fields per region, n = 3 animals per genotype). The scatter plots of in g-ratio is depicted on right for each region.

Indeed, transmission electron microscopy (TEM) images revealed a significant reduction in the number of myelinated axons in *Atm^-/-^* mice (Fig. 2G-I). Axon diameter, by contrast, was significantly increased in the CC and CBM, although not in the cortex. The g-ratio was largely unchanged in the *Atm^-/-^* brain, apart from a significant increase (thicker myelin) in the *Atm^-/-^*corpus callosum (Fig. 2I). Of note, the axonal membrane and the inner membrane of the myelin wrapping tended to be more consistently separated in the mutants (yellow arrowheads, Fig. 2H). Consistent with the preferential loss of MAG protein (Fig. 2C-E), this suggests a difference in the strength of the interaction between neurons and oligodendrocytes and MAG is a glycoprotein found in the initial wraps of myelin that are immediately adjacent to the axon, while MOG is a glycoprotein found on the outer surface of the myelin ensheathment; MBP and PLP are found in the lipid-rich dense bands of the compact myelin (Fig. 2F). Together, this observation suggested that ATM deficiency primarily interferes with the interaction between neurons and OLs. Their close apposition is lost while other structural and biochemical features of myelin, though quantitatively reduced, are largely maintained in *Atm^-/-^* animals.

### Oligodendrocyte pathology in Atm-knockout mice

To learn the progression of myelin defects over time, we compared the OL populations in wildtype and *Atm^-/-^* at 1 and 6 MO by immunohistochemistry and found region-specific changes of OL population using Olig2 as the pan-OL marker, NG2 as the OPC marker and MyRF/CC1 as the mOL markers (Fig. 3A-E).

**Figure 3.**
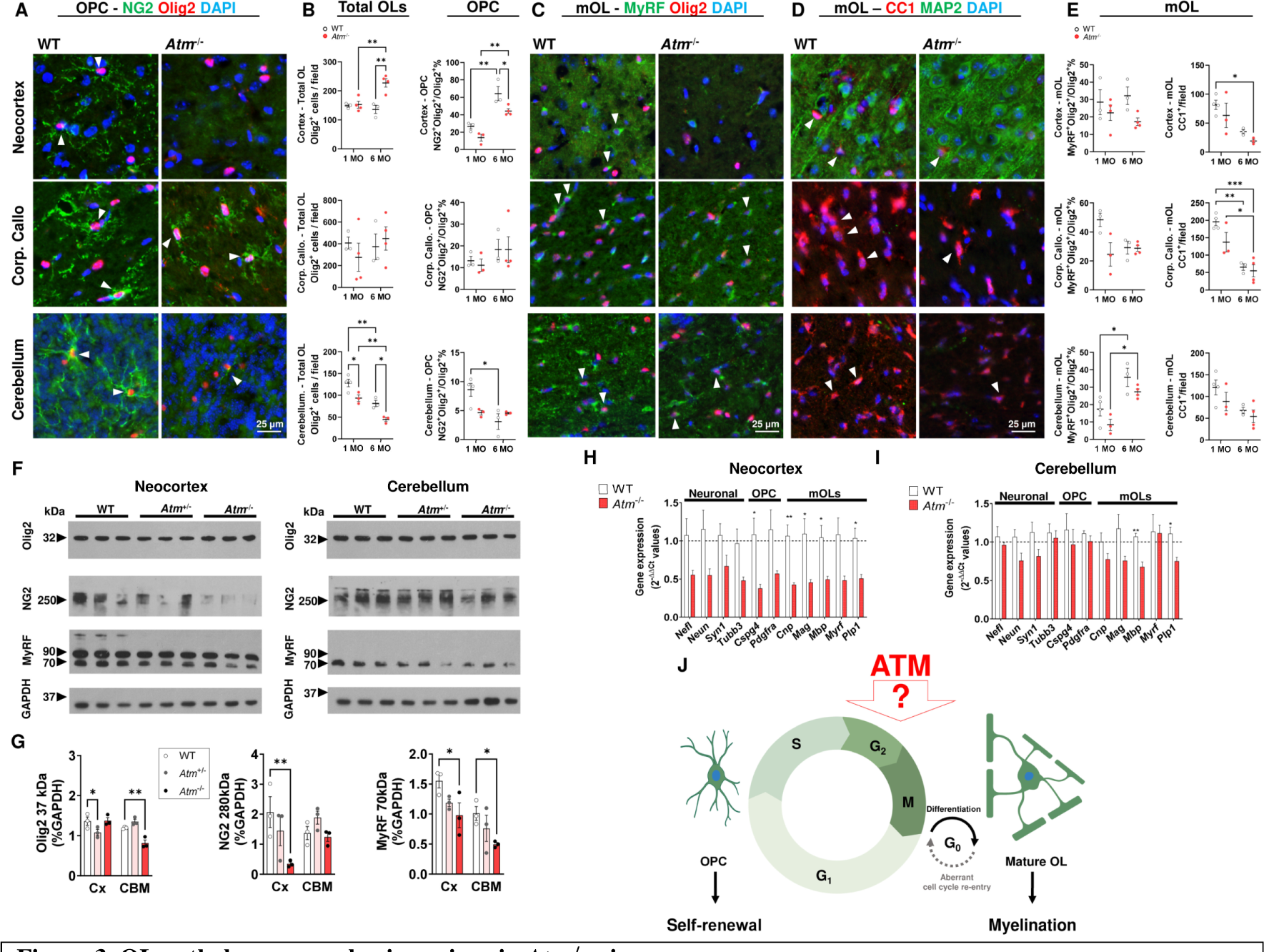
OL pathology across brain regions in *Atm*-/- mice **A** Representative immunohistochemistry image showing NG2+ (green) Olig2+ (red) OPC (filled arrowheads) in cortex, corpus callosum and cerebellum of the *Atm*-/- mouse brain at 1 month of age. **B** Quantification of total OL (Olig2+) density and NG2+Olig2+ OPCs proportion showed a differential age- and genotype-dependent change of OLs and OPCs across the neocortex and cerebellum of *Atm*-/- mouse at 1 and 6 months of age (MO). Except for cerebellar OPCs, genotype and aging contributed to a significant effect to both cell types (p < 0.01) but no significant change was found in the corpus callosum. Representative immunohistochemistry images showing myelinating mOL (MyRF+Olig2+ double-positive or CC1+ cells) were shown in **C** and **D**, respectively. **E, F** Quantification of MyRF+(green) Olig2+(red) mOL and CC1+ mOL fraction proportion showed that *Atm*-/- genotype contributed to a significant reduction of MyRF+ Olig2+ among all regions (p < 0.05) while aging is associated with the loss of CC1+ mOL in the 6 MO mice (p < 0.05), pairwise differences were depicted on graphs (Two-way ANOVA with Holm-Šídák’s multiple comparisons test; *p < 0.05, **p < 0.01, ***p < 0.001). **F** Immunoblots of OL proteins (Olig2, NG2, and MyRF) in the neocortex and cerebellum showing *Atm*-/- genotype is associated with a major change of MyRF (p = 0.0084) and NG2 (p = 0.0247), but not Olig2 (p = 0.0722), with quantification against GAPDH as shown in **G** (Two-way ANOVA with Holm-Šídák’s multiple comparisons test; * p < 0.05, **p < 0.01). **H, I** The expression of cell type-specific genes as measured by real time PCR in lysates of neocortex and cerebellum (unpaired t-test, \**p* < 0.05, \*\**p* < 0.01). **J** We hypothesize that ATM functions are required for the regulation of the cell cycle progression in self-renewing OPC and post-mitotic mOL to maintain myelination.

In the CX, the pan-Olig2^+^ OL density remained constant in wildtype, but it was unexpectedly increased by 67% in *Atm^-/-^* at 6 MO (P = 0.0039). The cortical NG2^+^ OPC fraction significantly increased with age in both genotypes (P = 0.0001), but it was significantly lower in *Atm^-/-^* mice by more than one-third (P = 0.0069). These age-related increases of cortical OL and OPC in *Atm^-/-^* CX were not accompanied by a corresponding increase of MyRF^+^ or CC1^+^ mOL. Instead, *Atm^-/-^* genotype contributed to a significantly lower MyRF^+^ cell number in all ages (P = 0.049), and the lowest number of CC1^+^ mOL at 6 MO. In CC, the variability in cell density was marked for OL at all stages of maturation, with no significant differences of the total Olig2^+^ OL or NG2^+^ OPC population being observed. However, a significant decrease in the density of fully mature CC1^+^ mOL was found in *Atm^-/-^* mice at 6 MO.

In the cerebellum, Olig2^+^ OL density (all stages of the OL lineage) was significantly lower in *Atm^-/-^* than wildtype and this deficit worsened with age – from 27% at 1 MO to 45% by 6 MO (P = 0.001). The OPC population (NG2^+^) was more than halved between 1 and 6 months in wildtype, but it remained constantly low in *Atm^-/-^* mice. In a similar fashion, cerebellar CC1^+^ mOL was reduced with aging in both strains. Intriguingly, despite the age-dependent decreases in most stages of the OL lineage, we observed an increase in the number of actively myelinating (MyRF^+^) cerebellar OL between 1 and 6 months in both genotypes (P = 0.0004).

As the myelin and OL changes were found in *Atm^-/-^* mice as early as one month of age, we confirmed such early region-specific changes at the protein level using immunoblotting and gene expression at this age (Fig. 3F-I). Consistent with the histological findings, Olig2 expression was significantly reduced in *Atm^-/-^* CBM by 31% compared with wildtype (P = 0.0063), but there were no changes in CX. NG2 expression was reduced by 83% in *Atm^-/-^* compared to wildtype (P = 0.0055). For MyRF, the trend towards a reduction that we observed with immunocytochemistry was significant with respect to the levels of total protein with a 37% reduction in CX (P = 0.0297) and a 51% reduction in CBM (P = 0.0498). At the gene expression level, OPC-associated genes (*Cspg4* [i.e., NG2]) and mOLs genes (*Mbp, Plp, Myrf* and *Mag*) were significantly downregulated in *Atm^-/-^* CX. In the cerebellum, despite the overall reduction of OLs and OPCs numbers, most myelin-related transcripts were largely unchanged in *Atm^-/-^* mice. These were degenerative, not developmental defects as there were no differences in Olig2^+^ cell numbers in any brain region of young postnatal mice (P4-P6) (Fig. S2). Together, these data suggested that OL differentiation has failed in the adult *Atm^-/-^* CX, while OL is compromised in the *Atm^-/-^* CBM (Fig. 3J).

### Cell cycle dysregulation in mOL of Atm-knockout mice

ATM repairs DNA damage and regulates cell cycle progression to prevent any cells with compromised genomic integrity from dividing. In *Atm*^-/-^ mice, normally post-mitotic cerebellar PC, re-enter a progression of cell cycle events and die (Kuljis et al., 1997; Lavin, 2013; Li et al., 2011; Xu et al., 1996; Yang and Herrup, 2005). To test if ATM similarly regulates cell cycle in OL, we first asked if post-mitotic mOLs also enter an abnormal cell cycle *Atm*^-/-^ mice. The re-expression of cyclin D1 protein in G1/G0 phase is a first step towards cell cycle re-entry-related cell death in neurons (Chauhan et al., 2020; Freeman et al., 1994; Kranenburg et al., 1996). Reproducing earlier findings, immunohistochemistry showed a robust re-expression of cyclin D1 in cerebellar neurons and added data that cell cycle re-entry was even more pronounced in cortical neurons (4.5 folds, P = 0.0001) and, to a lesser extent, in cerebellar neurons in *Atm^-/-^* mice at 1 MO (Fig. 4A, B). We extended our observations to later ages and found that neuronal ectopic cell cycle events were substantially diminished at 6 months of age.

**Figure 4.**
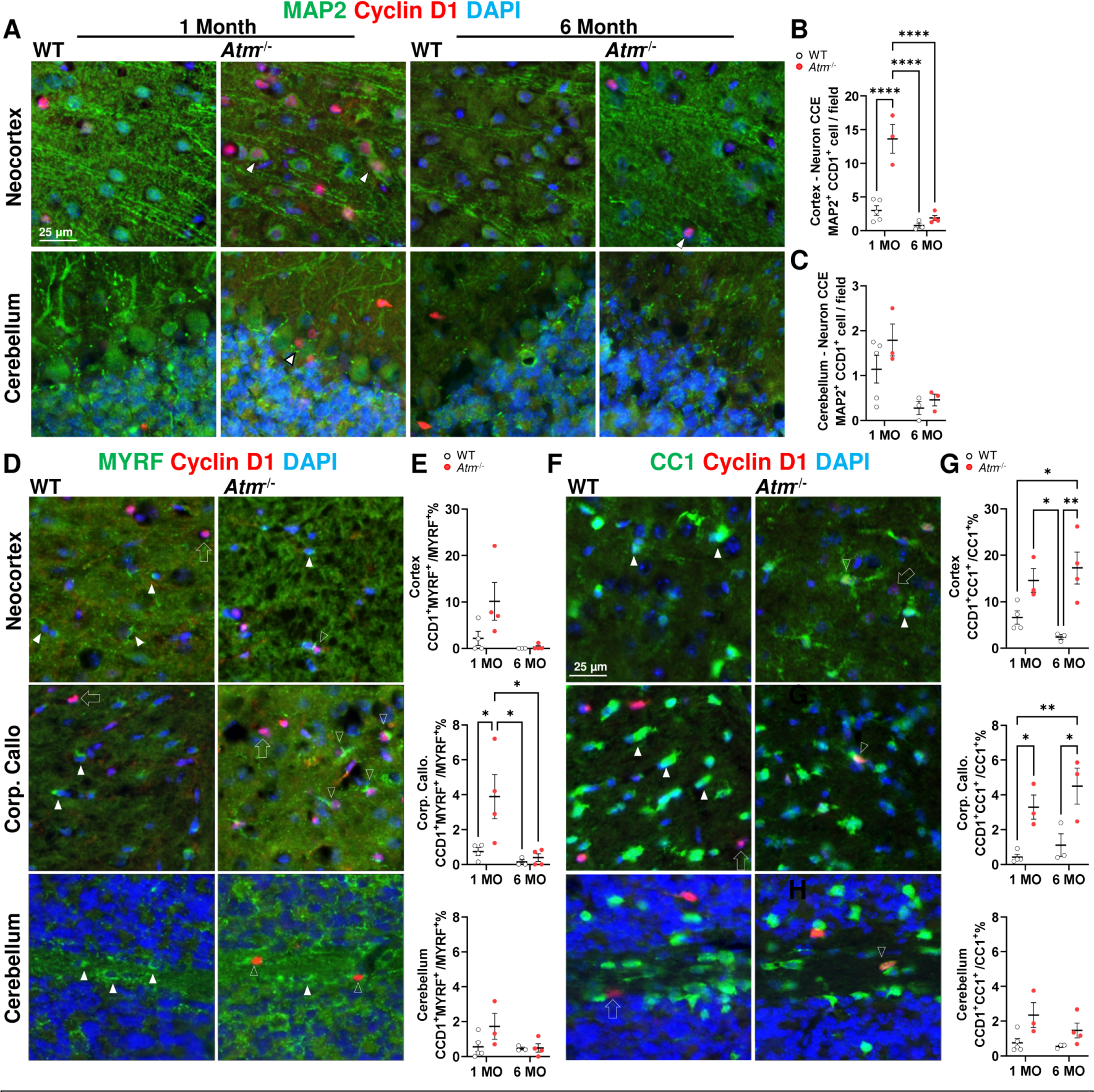
Aberrant cell cycle protein re-expression in neurons and mOL of *Atm*-/- mice **A** Representative image showing the early neuronal pathology in form of aberrant re-expression of the cell cycle-related protein, cyclin D1 (red; CCD1) in post-mitotic neurons (MAP2+, green) of the neocortex and cerebellum in *Atm*-/- mice at 1 and 6 MO (arrowheads). **B, C** Image analysis showed that *Atm*-/- genotype significantly increased aberrant cyclin D1 reexpression in cortical neurons (p < 0.0001) but not cerebellar neurons. Such event was abolished by aging in both regions (p < 0.0039). **D-H** Representative image aberrant re-expression cyclin D1 (red) was also found in **D** MyRF+ mOL (green) and **F** CC1+ mOL (green) across cortex, corpus callosum and cerebellum of the *Atm*-/- mouse at 1 month of age (MyRF+ or CC1+/cyclin D1+, open arrowhead; MyRF+ or CC1+/only, filled arrowhead; cyclin D1+ only, open arrows). **E** Quantifications showed that *Atm*-/- contributed to a significant change of abnormal MyRF+ cyclin D1+ mOL only in corpus callosum (p = 0.0355), but contributed to a significant change of abnormal CC1+ cyclin D1+ mOL in cortex (p = 0.0009), corpus callosum (p = 0.0009) and cerebellum (p = 0.0103) as shown in **G**. Statistical analysis by two-way ANOVA with Holm-Šídák’s multiple comparisons test; * p < 0.05, **p < 0.01.

We extended these histology findings in neurons to the post-mitotic myelinating mOLs of *Atm*^-/-^ mouse brain (Fig. 4D, E). As with the neurons, the abnormal cyclin D1 re-expression was found in actively myelinating MyRF^+^ mOLs in the CC of *Atm^-/-^* mice at 1 MO (P = 0.0355). The cyclin D1^+^ MyRF^+^ double positive cells were five-fold higher than in wildtype (Fig. 4F, G). Despite the robust increase of cyclin D1^+^ MyRF^+^ in CX and CBM, no statistical significance was reached. Intriguingly, in both *Atm^-/-^* and wild type animals, cell cycle activity in MyRF^+^ cells dropped nearly to zero by 6 months of age. We next examined the cell cycle activity in the entire CC1^+^ population of mOL. The number of cyclin D1^+^ CC1^+^ mOL was significantly increased in the *Atm^-/-^*mice across the CX (>2.2-fold, P = 0.0009), CC (>4.1-fold, P = 0.0009) and CBM (>2.7-fold, P = 0.0103). The increase was found at both ages examined (Fig. 4G) and this pattern of cell cycle activity closely tracked the pattern of mOL loss in the *Atm*^-/-^ mice (Fig. 3).

### OPC cell cycle progression is vulnerable to DNA damage and ATM deficiency

As the abnormal cell cycle events in mOL coincided with the appearance of neuronal deficits in *Atm*^-/-^ mice (Fig. 4A, Fig. S3A-C), these data alone are insufficient to answer the question whether cell cycle dysregulation is a cell autonomous consequence of ATM deficiency in the OL themselves or whether they are responding indirectly to a preceding neuronal pathology. While the OPC divides to maintain the pool of progenitors needed for myelin plasticity, cell cycle machinery may be equally important in the post-mitotic mOL (Herrup, 2013; Katsel et al., 2008). We propose that ATM is likely to regulate both ends of the OL life cycle. To test this idea, we turned to cell culture using both primary OL cultures as well as the OPC cell line, Oli-Neu (Biname et al., 2013).

In primary OPC culture from wildtype pups, endogenous ATM-IR (2C1, immunoreactivity) was identified in both nucleus and cytoplasm of the NG2^+^ cells; nuclear ATM appeared as multiple foci (Fig.5A). Activated ATM (pATM^ser1981^) was found located in the nucleus but was for more prominent in the cytoplasm. Cytoplasmic pATM^ser1981^-IR extended throughout the NG2^+^ cellular processes (Fig.5B, above). To control for non-specific antibody bindings, we applied a stable and highly selective ATM kinase inhibitor, KU-60019 (10 µM) (Golding et al., 2009; Kang et al., 2017; Khalil et al., 2012; White et al., 2008) but reduced the pATM^ser1981^-IR nearly to the background level (Fig.5B, below; Fig. 5C). KU-60019 had virtually no effect on the survival of either OPCs (NG2^+^) or total OLs (Olig2^+^). Inhibition of ATM activity increases DNA damage by slowing down genomic repair (Bourseguin et al., 2022; Chow et al., 2019a; Mehta and Haber, 2014; Woodbine et al., 2011). We confirmed this in the Oli-Neu and OPC cultures by demonstrating a KU-60019-mediated, dose-dependent accumulation of 53BP1^+^ and γH2AX^+^ foci (Fig. S4A-I). The OPC was also highly sensitive to hydrogen peroxide (H2O2)-mediated oxidative stress leading to 53BP1^+^ and γH2AX^+^ DSB foci formation (Fig. S4F-I), as reported earlier (Back et al., 2002; Bagi et al., 2018), and these oxidative stresses and DSBs were identified in the OL population in *Atm*^-/-^ mice (Fig S4J, K)

**Figure 5.**
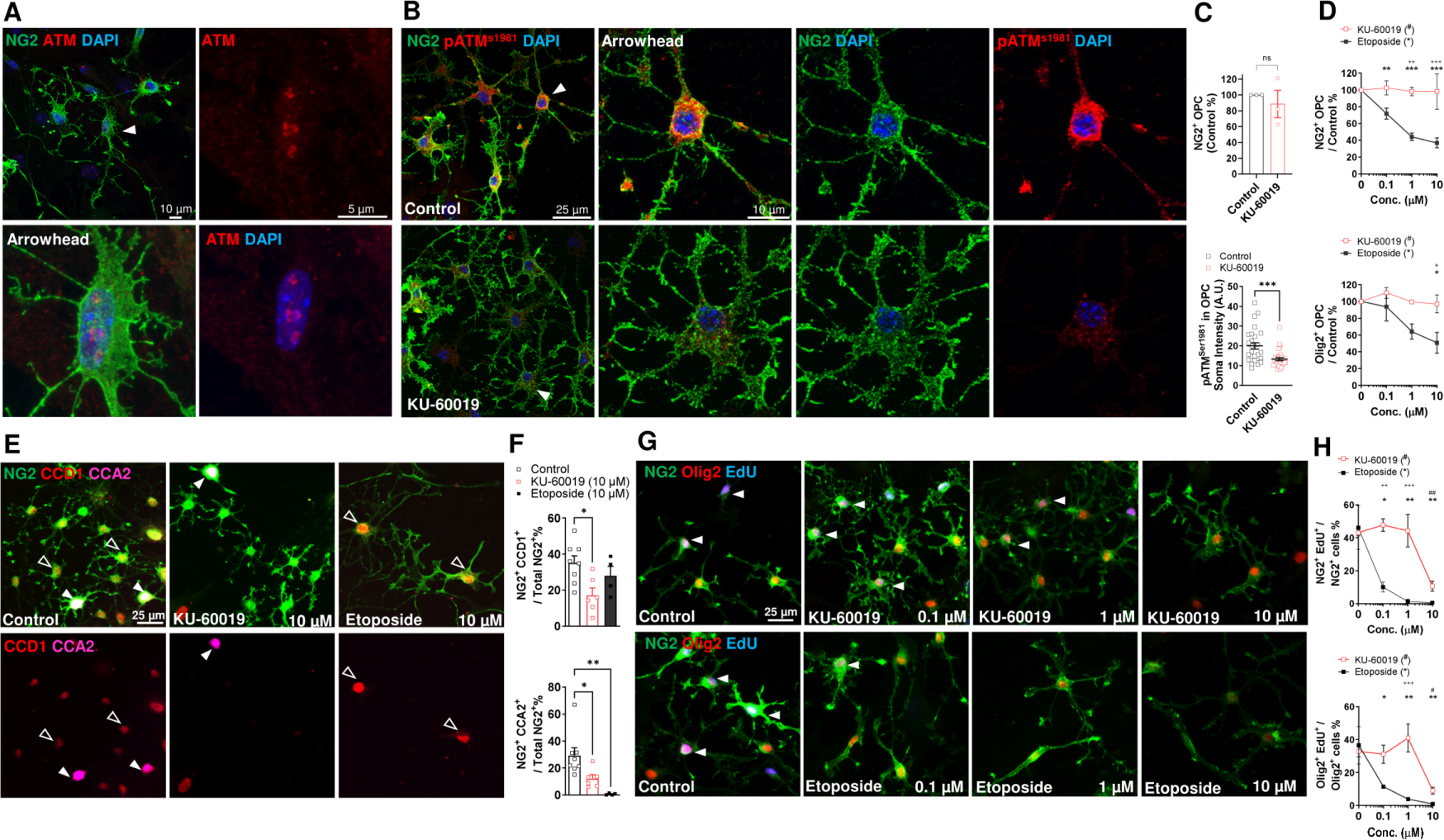
Functional ATM expression in cycling OPC A. Representative confocal image showing ATM-IR (red) in both nucleus and cytoplasm of cultured NG2+ OPC (green, arrowhead). **B** Activated ATM (pATMS1981, red) was detected in both nucleus and cytoplasm of the NG2+ OPC, but such level was attenuated by KU-60019 (10 μM, 24 h bottom) without affecting cell viability, as shown in **C**. **D** A dose response study of KU-60019-mediated ATM inhibition (red) and etoposide-mediated DSB-DNA damage (black, 0 – 10 μM, 24 h) on OPC density and Olig2+ OL density showing the OPC is highly sensitive to DNA damage but not ATM inhibition. **E, F** OPC cell cycle activity denoted by cyclin D1 (CCD1, red) and cyclin A1 (CCA2, magenta) were attenuated by KU-60019 and etoposide *in vitro*. **G** OPC (NG2+ green, Olig2+, red) cell cycle progression towards S phase was measured by EdU incorporation revealed by ClickIT chemistry (blue) in the presence of increasing doses of KU-60019 and etoposide as indicated**. H** Quantification of EdU assay showing the entry of S phase is highly sensitive to DSB and sensitive to high level of ATM inhibition (*etoposide vs control #KU-60019 vs control, +KU-60019 vs etoposide, one-way ANOVA with Dunnett’s multiple comparisons test or */*#/+p* < 0.05, **/#*#/++p* < 0.01, ***/##*#/+++p* < 0.001, n = 3 - 4).

We next asked if etoposide, a selective topoisomerase II inhibitor that induces DSB formation, triggered a similar dose-dependent effect. We found that etoposide was toxic to cells of OL lineage (Fig. 5D). As both DSB formation and ATM activity are known to alter cell cycle kinetics, we used cyclin D1, cyclin A2 and EdU incorporation to study the effect of the two compounds on OPC cell cycle (Fig. 5E-H). Despite the lack of effects on the OPCs number, KU-60019 reduced the cyclin D1^+^ and cyclin A2^+^ OPC by more than half (Fig. 5F); etoposide had a similar effect on cyclin D1 (a G1 phase marker), but virtually abolished cyclin A2 (a S-phase marker). The strong S-phase suppression was validated by the near total absence of EdU uptake at an etoposide concentration (0.1 µM) 100-fold lower than KU-60019 (Fig. 5H).

### OPC differentiation requires ATM activity

The cytoplasmic expression of pATM^ser1981^ in OPC prompted us to investigate the functions of ATM in OL outside the nucleus. In wildtype OL culture, nuclear and cytoplasmic pATM^ser1981^-IR were identified in the newly formed MBP-expressing mOL (Fig. 6A). We are particularly intrigued by how the cytoplasmic pATM^ser1981^ localized not only inside the cell body, but also along the many MBP^+^ cellular processes (Fig. 6A, arrowheads, right panel) and how, despite the proximity, the majority of pATM^ser1981^- and MBP-IR did not co-localize. In fully mature OL, pATM^ser1981^-IR persisted in the major branches of cytoplasm but remained absent in the vallate of MBP^+^ myelin sheets (Fig. 6B, arrowheads, right panel).

**Figure 6.**
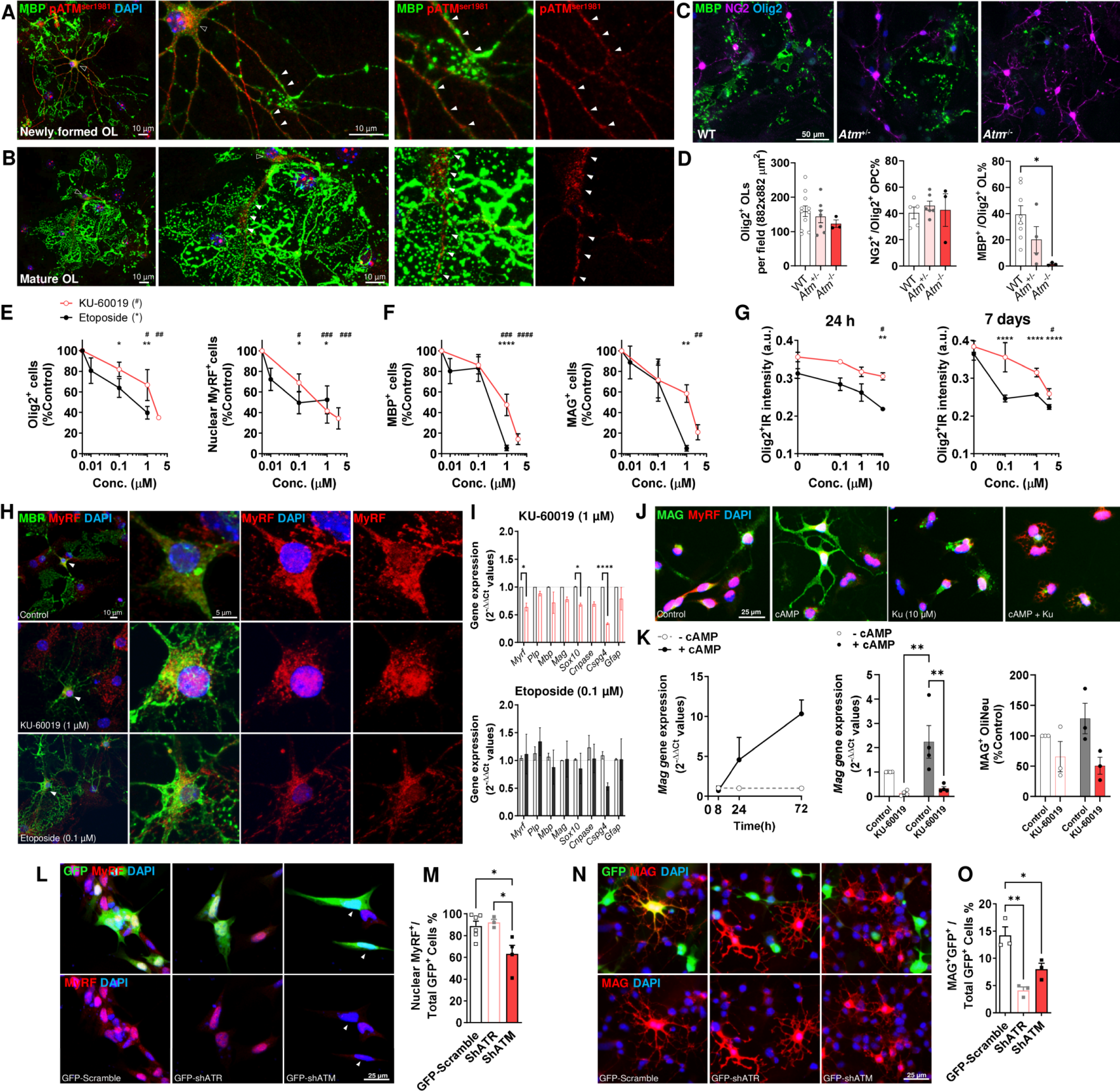
Functional ATM expression in differentiating OPC A. *Left* Representative confocal image showing activated ATM (pATMS1981, red) was detected in the nucleus (open arrowhead) and cytoplasm of newly formed MBP+ OLs (green). *Middle* Higher magnification showed pATMS1981+ in the nucleus and in the cytoplasmic branches (filled arrowheads). *Right* The cytoplasmic pATMS1981 were found in close proximity to the developing myelin sheet but did not colocalize with MBP. **B** *Left* Representative confocal image showing activated ATM (pATMS1981, red) was similarly detected in a well-established mOL with elaborated myelin sheet. *Middle* the pATMS1981 was detected as nuclear foci and along the major cytoplasmic branches harboring the myelin sheet *right* **C, D** In *Atm*-/- OL culture, Olig2+ OL and NG2+ OPC were not affected by Atm-deficiency, but the formation of MBP+ mOL was reduced in a gene-dose dependent fashion in *Atm+*/- and *Atm*-/- culture, suggestive of differentiation failure. **E-G** Results of a dose response study of KU-60019-mediated ATM inhibition (red, 0.1 – 2.5 μM) and etoposide-mediated DSB-DNA damage (black, 0.01 – 1 μM) throughout a 7 DIV differentiation period of OPC culture. **E** Both ATM inhibition and DSB induction significantly reduced the number of Olig2+ OL and MyRF+ mOL in a concentration-dependent fashion. **F** Midto- high dosage of KU-60019 and etoposide nearly abolished the formation of MBP+ and MAG+ mOL. **G** Both KU-60019 and etoposide treatment gradually (24 h and 7 days treatment) reduced the intensity of Olig2-IR in the surviving OL population in a concentration-dependent fashion. (*etoposide vs control, #KU-60019 vs control, one-way ANOVA with Dunnett’s multiple comparisons test or */*#p* < 0.05, **/#*#p* < 0.01, ***/##*#p* < 0.001, ****/###*#p* < 0.0001, n = 3 - 4). **H** Confocal images showing nuclear MyRF localization in mOL after chronic incubation with KU-60019 (1 μM) and etoposide (0.1 μM). **I** KU60019-, but not etoposide-mediated reduction of Olig2 and MyRF, the two OL-specifying transcription was associated with a significant decline of myelin/OL gene transcription. **J** Immunocytochemistry of Oli-Neu differentiated with cAMP ± KU-60019 **K** cAMP-induced differentiation triggered a significant *Mag* transcription Oli- Neu, but pre-treatment of KU-60019 significantly reduced *Mag* expression (p = 0.0014) and MAG+ cell formation (p =0.0185). Two-way ANOVA with Holm-Šídák’s multiple comparisons test; * p < 0.05, **p < 0.01. **L, M** The use of shRNAmediated knockdown (green, GFP-sh) of *Atm* but not *Atr* significantly reduced the percentage of nuclear MyRF+ Oli-Neu. **N, O** shRNA-mediated knockdown (green, GFP-sh) of *Atm* and *Atr* significantly reduced MAG+ Oli-Neu formation. Oneway ANOVA with Tukey’s multiple comparisons test or \**p* < 0.05, \*\**p* < 0.01.

In *Atm*^-/-^ primary OL culture, the densities of Olig2^+^ OL and NG2^+^ OPC were not notably different from wildtype cultures (Fig. 6C), but the formation of MBP^+^ mOL was nearly abolished (reduced by 96%, p = 0.024) (Fig. 6D). To confirm that this effect was due to the loss ATM kinase activity, OPCs were incubated with low concentrations of KU-60019 (0.1, 1 and 2.5 µM) or etoposide (0.01, 0.1 and 1 µM) throughout the seven-day differentiation protocol. At the end of the incubation, both KU-60019 and etoposide significantly reduced the OL-specific transcription factor Olig2 and MyRF-expressing cells in a concentration-dependent fashion (Fig. 6E). KU-60019 (1 µM) also reduced MBP^+^ and MAG^+^ mOL formation by 52.5% (p = 0.0002) and 41.6% (p = 0.07), respectively, while etoposide (1 µM), reduced MBP^+^ and MAG^+^ mOL formation by 94.4% (p < 0.0001) and 94.7% (p = 0.0011) (Fig. 6F). As Olig2 and MyRF coordinate myelin transcription (Bujalka et al., 2013; Mei et al., 2013; Wegener et al., 2015), these results suggest that the OPC differentiation program is highly vulnerable to ATM inhibition and/or DNA damage. Indeed, the expression level of nuclear Olig2 in the surviving OLs was significantly reduced in OPC culture after acute (24 h, Fig. 6G, left) or chronic (7 days Fig. 6G, right) incubation with etoposide or KU-60019. In the surviving mOL, although the nuclear translocation of MyRF was unaffected by the treatments (Fig. 6H), KU-60019 treatment, but not etoposide, contributed to the overall reduction of OL gene transcription (p < 0.0001) where *Myrf* was reduced by more than 36% (Fig. 6I).

To define the additional roles for ATM kinase activity during OL differentiation, we turned to Oli-Neu, a murine OL cell line free from other glial cell types. Oli-Neu cells constitutively express nuclear MyRF and can be differentiated with cAMP into MAG-expressing mOL phenotype (Fig. 6J). Treatment of Oli-Neu with KU-60019 (10 µM) prior to cAMP induction reduced *Mag* gene transcription (p = 0.0014) and the number of MAG^+^ Oli-Neu (p = 0.0185, Fig. 6K). Finally, shRNA-mediated knockdown of ATM but not the related phosphatidyl-inositol 3-kinase family member, ATR (ATM and RAD3-related kinase) reduced the number of Oli-Neu with nuclear MyRF (Fig. 6L, M). As both shATM and shATR significantly blocked Oli-Neu differentiation by more than 50% (Fig. 6N, O), these data suggest that nuclear localization of MyRF requires ATM while myelin formation requires both ATM and/or ATR Together, these data suggested that ATM activity is not only important for cell cycle regulation of OPC but also for their differentiation program.

### ATM activity is also requited for cell cycle control in mature oligodendrocyte

To study ATM-mediated regulation of cell cycle in myelinating cells, we cultured mOL from wildtype pups and found the distribution of ATM and pATM^ser1981^ to be similar to that found in OPCs (Fig. 5A). In MBP-expressing mOL, ATM protein was located in both cytoplasm and in nuclear foci (Fig. 7A). Activated ATM (pATM^ser1981^)-IR was also found in the nucleus, soma and cytoplasmic processes of MBP^+^ mOL (Fig. 7B), but again not colocalized with the MBP^+^ myelin sheets (Fig. 7B, right). As KU-60019-mediated inhibition significantly reduced such pATM^ser1981^-IR (10 µM, 24 h) in the nucleus, soma, and the cytoplasmic processes of the mOL (Fig. 7C), endogenous ATM activity is present in the fully differentiated mOL.

**Figure 7.**
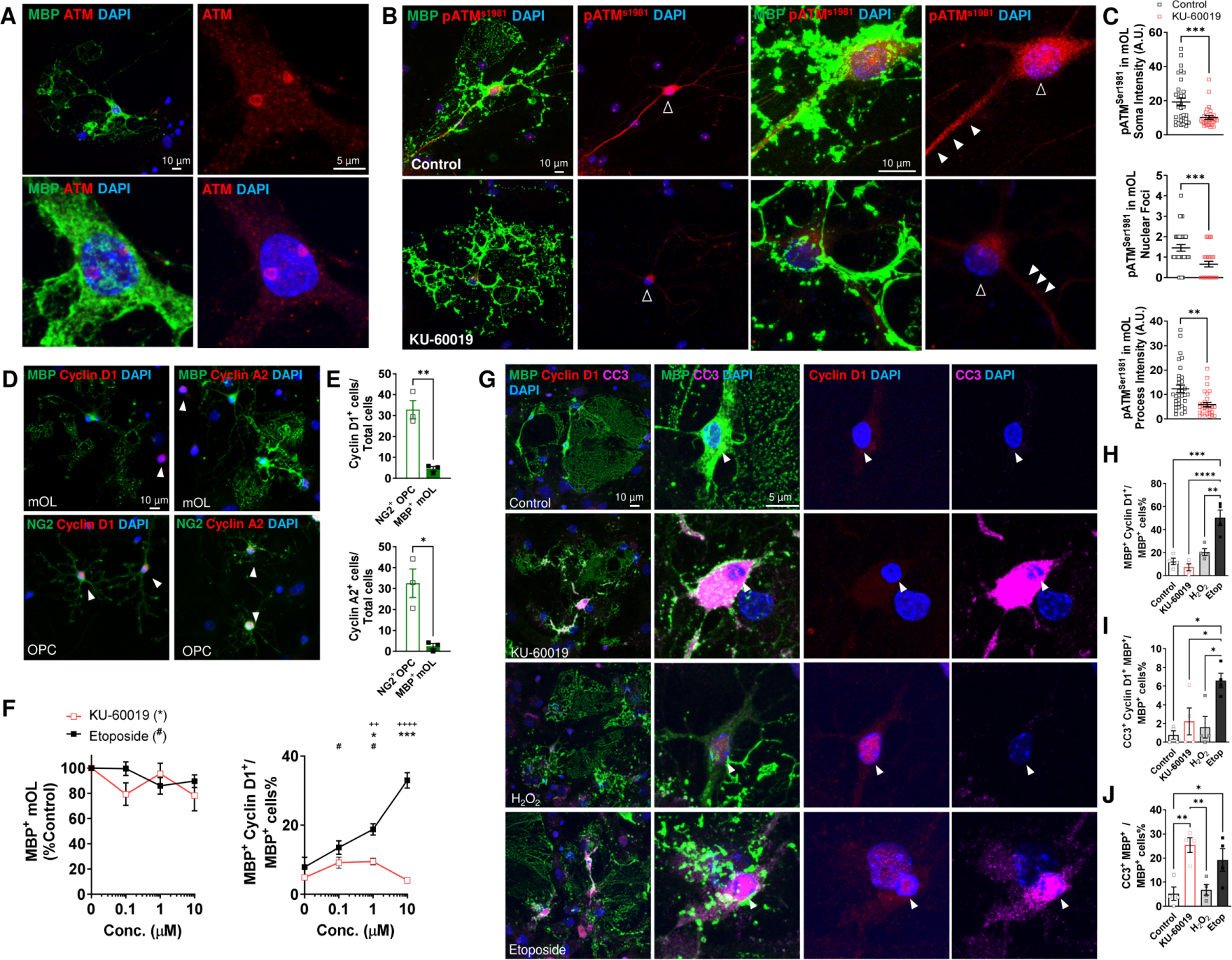
Functional ATM expression in myelinating mOL A. Representative confocal image showing ATM-IR (red) in both nucleus and cytoplasm of cultured MBP+ mOL (green). **B, C** *top* Activated ATM (pATMS1981, red) was detected in both nucleus, soma (open arrowheads) and cytoplasmic processes (filled arrowheads) of the MBP+ mOL. *bottom* The level was significantly reduced by KU-60019 (10 μM, 24 h). **D, E** Representative immunocytochemistry showing cyclin D1 and cyclin A2-expressing NG2+OPC and MBP+ mOL in culture at DIV7. The majority of cycling cells were OPC and confirms the postmitotic status of mOL. **F** A dose response study of KU-60019-mediated ATM inhibition (red) and etoposide-mediated DSB-DNA damage (black, 0 – 10 μM, 24 h) on cyclin D1 re-expression in mOL. etoposide at any concentrations and KU-60019 at low concentration significantly induced abnormal cyclin D1 expression in mOL, but the density of mOL was not affected. (*etoposide vs control #KU-60019 vs control, +KU-60019 vs etoposide, one-way ANOVA with Dunnett’s multiple comparisons test or two-way ANOVA with Holm-Šídák’s multiple comparisons test */*#p* < 0.05, *++p* < 0.01, \*\*\**/+++p* < 0.001, n = 3 - 4). **G** Representative confocal images of cultured mOL (MBP+ green) pretreated for 24 h with KU-60019 (10 μM), H2O2 (50 μM) and etoposide (10 μM). Cell cycle activity was measured by immunolabelling for cyclin D1 (red); cell death was measured by the presence of cleaved caspase-3 (CC3, magenta). **H-J** Quantification of the percentage of cycling mOLs (**H** MBP+ CyclinD1+), dying mOLs with cell cycle event (**I** MBP+ CyclinD1+CC3+), and all dying mOLs (**J** MBP+ CC3+) mOLs showed that only DSB induced a significant cell cycle-related cell death in mOL while ATM inhibition induced a significant apoptosis independent of cell cycle events. One-way ANOVA with Tukey’s multiple comparisons test or \**p* < 0.05, \*\**p* < 0.01, \*\*\**p* < 0.001, \*\*\*\**p* < 0.0001.

In the wildtype mOL cultures at 7 DIV, most cells expressing cyclin D1 (G1, 32.8%) and cyclin A2 (G1/S, 32.5%) were NG2^+^ OPC. Only a small fraction of differentiated MBP^+^ mOL expressed cyclin D1 (4.5%) and cyclin A2 (2.4%), and suggestive of cell cycle exit upon their successful differentiation (Fig. 7D, E). To test if ATM inhibition or accrued DNA damage or both drive the abnormal cell cycle- protein re-expression observed in the *Atm*^-/-^ mice, we tested the effect of KU-60019 or etoposide (0.1 to 10 µM, 24 h) on mOL at 7 DIV. Unlike OPC culture, the number of MBP^+^ mOLs were not sensitive to KU-60019 or etoposide at any concentration (Fig. 7F). However, a significant re-expression of cyclin D1 was found in up to 32.9% of the MBP^+^ mOLs upon etoposide application at any concentration, while KU-60019 treatment at low concentrations increased the cyclin D1^+^ MBP^+^ mOL by 9.5% (0.1 and 1 µM, 24 h). In this range of concentration, neither treatment had a significant effect on the number or cell cycle activity of primary embryonic cortical neurons (Fig. S3D) and suggested that mOL may be more sensitive to both DNA damage and ATM inhibition than neurons.

Aberrant cell cycle progression in mOL is associated with cell death (Tse et al., 2018). Using cleaved caspase 3 (CC3) as a measurement of apoptosis, we compared the effect of KU-60019 (10 µM), H2O2 (50 µM), and etoposide (10 µM, all 24 h, Fig. 7G). Etoposide was the only treatment that induced a significant re-expression of cyclin D1 (cyclin D1^+^ MBP^+^, 50.2%) and ectopic cell cycle-related cell death (cyclin D1^+^ CC3^+^ MBP^+^, 6.6%) in MBP^+^ mOLs (Fig. 7H, I). In contrast, KU-60019 directly triggered apoptosis 25.3% of the mOLs population without inducing cyclin D1 (Fig. 7J). Curiously, unlike the NG2^+^ OPC population, oxidative stress induced by H2O2 had no toxic effects on mOLs.

### ATM regulates DSB-induced cell cycle-related cell death in mature oligodendrocytes

Our experiments thus far show that DSB-DNA damage, and thereby ATM activation, significantly reduced cell cycle progression in self-renewing OPC and cause aberrant cell cycle-associated apoptosis in post-mitotic mOL. In contrast, ATM inhibition also significantly reduces cell cycle progression in OPC, while causing cell cycle-independent apoptosis in mOL (Fig. 5, Fig. 7). To clarify the difference between naturally occurring cell division and in DSB-induced cell cycle re-entry process in mOL, we investigated cell cycle progression of fully established mOL culture (14 DIV) using OL-specific growth factors (PDGF-AA and bFGF, 10 ng/mL), KU-60019 (10 µM) or etoposide (10 µM). We measured cyclin D1 expression as a measure of cells in G1 phase, cyclin A2 to measure cells at the G1/S transition and EdU incorporation as a marker for cells in S phase. Cell death (apoptosis) was measured with cleaved caspase-3 (CC3) (Fig. 8A-E).

**Figure 8.**
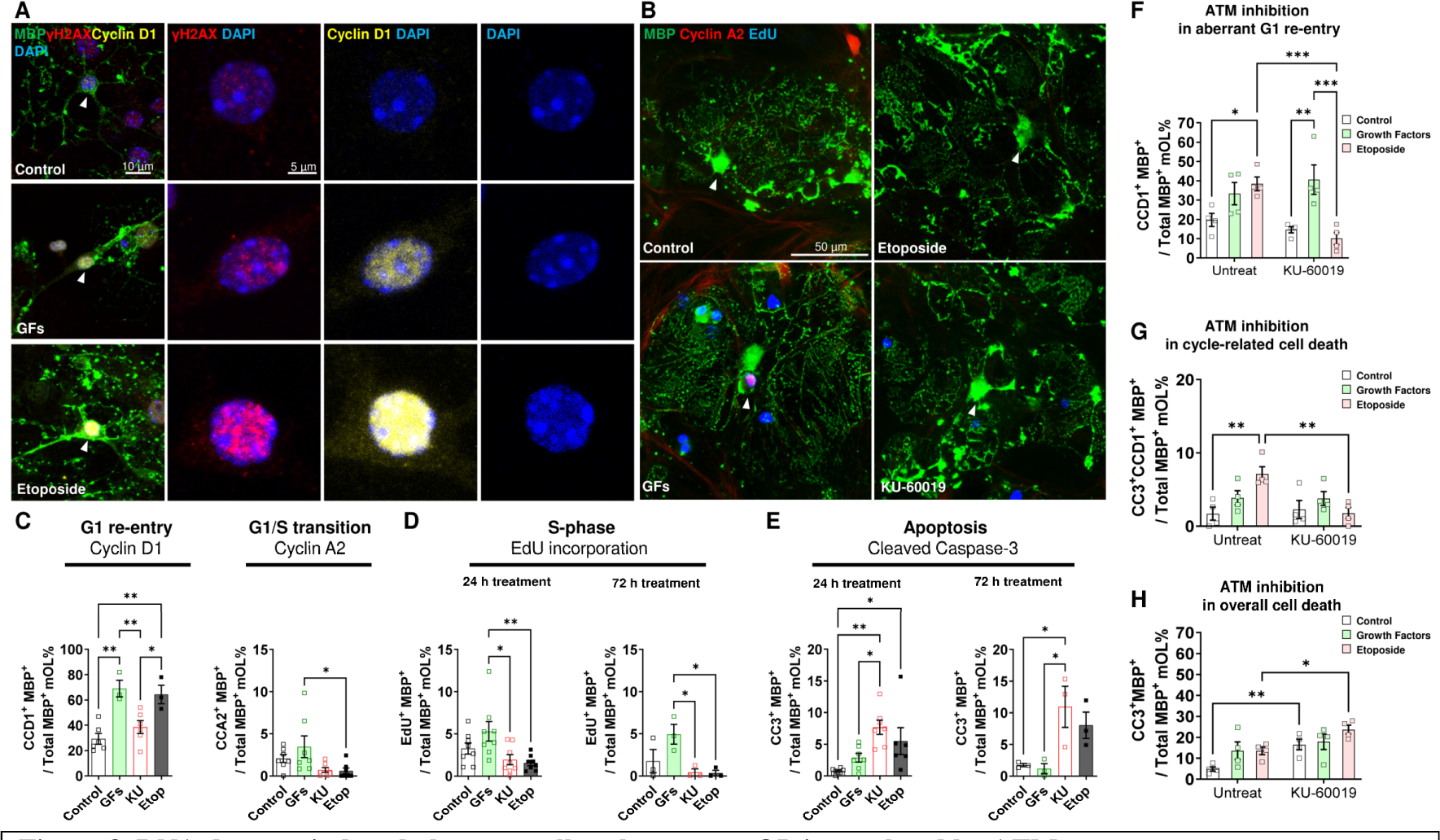
DNA damage-induced aberrant cell cycle events mOL is regulated by ATM A. Confocal image showing aberrant cell cycle events in mOL were induced by etoposide or OL-specific growth factors (GFs, PDGF-AA and bFGF, 2 ng/mL) at 14 DIV (cyclin D1, yellow; γH2AX, red; and MBP, green). **B** Confocal image of EdU incorporation assay (Blue) and cyclin A2 (red) immunocytochemistry showing the abnormal re-entry of MBP+ mOL (green) into the G1/S transition phase of the cell cycle is only induced by growth factors. **C** Quantification showed that growth factors (PDGF-AA and bFGF, 2 ng/mL) and etoposide (10 μM), but not KU-60019 (10 μM) significantly induced cyclin D1 re-expression in MBP+ mOL. No treatment significantly induced cyclin A2 re-expression. **D** Quantification showed only growth factors increased the number of EdU positive mOL after 24 and 72 hours of incubation but did not reach statistical significance. **E** etoposide and KU-60019 (10 μM), but not growth factor, significantly induced mOL apoptosis after 24 and 72 hours of treatment. One-way ANOVA with Tukey’s multiple comparisons test or \**p* < 0.05, \*\**p* < 0.01. **F-H** To investigate the role of ATM in regulating aberrant cell cycle and cell death in mOL, cultured mOL at 14 DIV was treated with growth factors (PDGF-AA and bFGF, 2 ng/mL) and etoposide (10 μM) for 24 hours with or without the pre-treatment of KU-60019 (10 μM). **F** etoposide-induced, but not growth factors-induced, aberrant cyclin D1 reexpression in mOL was abolished by prior ATM inhibition. **G** Similarly, etoposide-induced aberrant cell cycle-related cell death was abolished by prior ATM inhibition. **H** Cleaved caspase 3 immunocytochemistry showed that KU-60019-mediated ATM inhibition induced a significant increase of mOL apoptosis, and such apoptosis was potentiated in the presence of etoposide but not growth factor treatment. Statistical analysis by two-way ANOVA with Holm-Šídák’s multiple comparisons test; * p < 0.05, **p < 0.01, ***p < 0.001.

In MBP^+^ mOL, growth factors drove the abnormal re-expression of cyclin D1 with a significant increase of DSB (γH2AX^+^ foci) in the nucleus after 24 hours treatment (64%), similar to those found after etoposide exposure (69%, Fig. 8C). However, neither growth factors nor etoposide significantly increased the cyclin A2^+^ MBP^+^ mOL after 24 hours. KU-60019-inhibited ATM expression had no effects on cyclin D1 or cyclin A2. To study S phase, EdU was applied to mOL 30 minutes prior to the treatments. Although growth factors slightly increased the number of EdU^+^MBP^+^ mOL, it did not reach statistical significance even after 72-hours (Fig. 8B, D). Intriguingly, etoposide and KU-60019 appeared to suppress the background levels of cyclin A2 expression and EdU incorporation. For apoptosis, both KU-60019 and etoposide induced a significant increase of CC3^+^ apoptotic mOL, but growth factors had no effects even after 72 hours incubation (Fig. 6E). Together, while both growth factors and DSB could similarly induce cyclin D1 re-expression in mOL, only DSBs were associated withcell cycle-related death. Importantly, none of the treatments drove these post-mitotic cells beyond the G1 phase.

Cyclin D1 is not only associated with cell cycle check point control at G1, but is also associated with apoptosis in post-mitotic cells (Freeman et al., 1994; Kranenburg et al., 1996) and with DNA repair mechanism (Jirawatnotai et al., 2011; Rivellini et al., 2022; Shimura et al., 2014; Zezula et al., 2001). As a test, we asked if the growth factor-induced and DSB-induced cyclin D1 re-expression in mOL was an ATM-regulated mechanism controlling cell cycle progression and cell death. Cultures of mOLs were treated with KU-60019 (10 µM), 30 min prior to the addition of growth factors or etoposide. We found that KU-60019 pre-treatment completely suppressed the DSBs-induced, but not growth factor-induced, re-expression of cyclin D1 (p = 0.0004, -73.6%, Fig. 8F). Second, KU-60019 pre-treatment also abolished cell cycle related cell death (CC3^+^ cyclin D1^+^ MBP-expressing mOL; p = 0.0011, Fig. 8G). Third, while KU-60019 pre-treatment induced apoptosis in mOL, it also potentiated the cell death mediated by DSB (+74%, p = 0.02), but not by growth factors (Fig. 8H). Finally, despite the significant induction of cyclin D1 re-expression, growth factors neither mediated a significant apoptosis nor were affected by the absence of ATM activity. Together, these findings suggested that ATM activity is required to regulate the DSBs-induced, but not growth factor-induced, cyclin D1 re-expression and the associated cell death in postmitotic mOLs.

## DISCUSSION

Myelin deficits of the central nervous system are a prominent pathological feature of A-T (Chung et al., 1994; Sahama et al., 2015; Sahama et al., 2014a; Sahama et al., 2014b). While these abnormalities have been assumed to be secondary consequences of the pronounced neuronal degeneration, direct evidence for this assumption has been lacking. Indeed, WM degeneration and possible abnormal myelin turnover are seen early in the A-T disease process (Dineen et al., 2020; Sahama et al., 2015; Sahama et al., 2014a), a finding in agreement with the reduced number of Olig2^+^ and CC1^+^ cells found in our youngest case (16 years). At all ages, the numbers of OLs remaining in the cerebellum were not correlated with any measure of Purkinje cell integrity – cell number or cell size. If the OL dystrophy were only secondary to a neuronal phenotype, this would unlikely be the case. The data suggest that the myelin pathology in A-T might be both a direct consequence of ATM deficiency in the OL lineage itself as well as the indirect consequence of the loss of the interaction with neurons (Lai et al., 2021).

The widespread myelin deficits in *Atm*^-/-^ mice at the transcriptional, translational and ultrastructural level – recapitulate most of the WM abnormalities found in human A-T cases (Aguilar et al., 1968; De Leon et al., 1976; Sourander et al., 1966; Strich, 1966; Terplan and Krauss, 1969). The mouse data also reveals important heterogeneity among different brain regions. The combined histological and molecular picture suggests that in cerebellum, the number of cells of the OL lineage is reduced in *Atm*^-/-^ animals, but the synthetic activity of each individual OL is increased. In neocortex, by contrast, OL cell number is largely unaffected, but the level of message and protein for a range of OL-related genes is depressed. Further, by comparing one-month with six-month animals, we find that it is the mature, myelin-forming OL population (MyRF^+^ and CC1^+^) that is the most vulnerable to ATM deficiency. These findings support our hypothesis that there are aspects of the OL pathology in A-T or *Atm*^-/-^ mice that are independent of neuronal pathology (Cheng et al., 2018; Jiang et al., 2015; Li et al., 2012; Yang and Herrup, 2005). Rigorous determination of the sequence of pathological events, specifically, whether the pattern of degeneration in the *Atm*^-/-^ brain starts in the neuron or starts in the oligodendrocyte remains a question for future investigations.

ATM is one of the three key DDR kinases that facilitates DSB repair by suppressing the cell cycle during the repair process. Our findings make it clear that this DDR function of ATM also applies to the OL lineage. Unlike neurons, the OL lineage maintains a mitotically active progenitor cell population of OPC in the adult brain (Psachoulia et al., 2009; Young et al., 2013). The survival and cell cycle progression of OPCs are highly sensitive to DSBs formation and ATM activity deficiency, a conclusion supported by our observation that the reduction of OPC density in the *Atm*^-/-^ neocortex and cerebellum appears to be the consequence of failed OPC self-renewal. This supposition agrees with earlier observations in Nbn^Cns-del^ mice, where OPCs also fail to proliferate in the absence of Nbn, an upstream regulator of ATM (Liu et al., 2014). It is thus appears that ATM plays an important role as a “guardian of the genome”(Shiloh, 2014), in oligodendrocytes as much as it does in neurons. Loss of ATM leads to a block in the self-renewal of OPC and in their subsequent differentiation program. After OL differentiation has begun, ATM deficiency would appear to be responsible for a region-specific loss of mature OL and their myelin. Hypomyelination is a regular phenotype of syndromes caused by genetic mutations that compromise DNA repair (Tse and Herrup, 2017). Thus, mutations in ATM (Ataxia-telangiectasia (Aguilar et al., 1968; De Leon et al., 1976; Sourander et al., 1966; Strich, 1966; Terplan and Krauss, 1969)), NBS1 (Nijmegen Breakage Syndrome (Maraschio et al., 2001)), and to a lesser extent the mutation of MRE11 (Ataxia telangiectasia-like disorder (Oba et al., 2010; Palmeri et al., 2013)) all result in myelin pathology.

We and others have demonstrated that when ATM activity is deficient, the reduced DNA repair capacity will often lead to DSB formation (Bourseguin et al., 2022; Chow et al., 2019a; Mehta and Haber, 2014). Precursors of OL express low levels of antioxidants and are thus poorly defended against oxidative stress (Back et al., 2002) and here we demonstrated that OPCs are also vulnerable to DSB formation. Mature, myelinating OLs, by contrast, are resistant to oxidative stress, but susceptible to DSB. It is instructive to compare the effects of DSB on the OL lineage with a second type of terminally differentiated brain cell, the neuron. Like neurons, myelinating, mOLs exit the cell cycle at the beginning of their final differentiation program (Emery et al., 2009; Zhang et al., 2014). The loss of ATM signaling in neuron results in aberrant cell cycle re-entry (Jiang et al., 2015; Li et al., 2012; Li et al., 2013; Shen et al., 2016; Yang and Herrup, 2005; Yang et al., 2014). A similar connection between cell cycle and cell death is also found among OL in AD (Tse et al., 2018). Here, our cell culture experiments demonstrate that DSB induces the re-expression of cyclin D1 and cell death in postmitotic mOLs – a process that is coordinated by ATM activity. In A-T and *Atm*^-/-^ animals, ATM deficiency and the accrued DSB formation may act in combination to cause cell cycle-dependent and -independent cell deaths of mOLs.

There are, however, distinct differences between the cell cycle/cell death relationship in proliferative cells and terminally differentiated cells like neurons and mOL. In proliferative cancer cells, cyclin D1 expression is part of the DNA repair mechanism upon DSB formation (Jirawatnotai et al., 2011). But in postmitotic neurons, the re-expression of cyclin D1, along with Cdk4, E2F1 and PCNA, is part of the DSB-induced cell death mechanism (Zhang et al., 2020). Most notably, these “cycling” neurons appear to survive *in vivo* for months if not longer after initiating a cell cycle or enter senescence (Chow et al., 2019b; Herrup and Yang, 2007). Here, we show that the “cycling” postmitotic mOLs with abnormal cyclin D1 expression are at risk for cell death, but their death follows quickly. Indeed, these “cycling” mOL are unable to pass the G1/S transition or enter S phase. Importantly, we showed that the loss of DNA integrity is the only pathway by which a mOL can be forced to enter an ectopic cell cycle-related cell death. While OL-specific mitogens also drive abnormal re-expression of cyclin D1 in postmitotic OL, they do not cause cell death. This suggests that, only following the DSBs activation does ATM regulate cell death, perhaps by altering the choice between DNA repair and apoptosis.

The accumulation of OPCs in neocortex despite the reduced present of myelin in *Atm*^-/-^ brain is consistent with the idea that the OL differentiation program is blocked by ATM loss. We confirmed such dependence on ATM by showing the failure of OPC differentiation in ATM-deficient OL culture by both genetic and pharmacological means. It remains an open question, however, whether the block to differentiation occurs in the nucleus or cytoplasm. ATM plays roles in the cell cytoplasm as well as its nucleus (Cheng et al., 2018; Chow et al., 2019a; Li et al., 2012). Cytoplasmic ATM is found in or on organelles, including mitochondria (Valentin-Vega et al., 2012) and synaptic vesicles (Cheng et al., 2021; Cheng et al., 2018; Vail et al., 2016), where it serves a variety of functions independent of DNA repair (Cheng et al., 2018; Lee et al., 2018; Zhang et al., 2018). In neuronal cell lines, cytoplasmic translocation of ATM is associated with terminal differentiation (Boehrs et al., 2007).

During the OL maturation process activated ATM (pATM^ser1981^) is a prominent feature of the developing cytoplasmic branches of the MBP-positive mOLs, but such cytoplasmic function of ATM in OL remains obscure. ATM deficiency inhibited OPC maturation despite the intact MyRF nuclear localization (Emery et al., 2009). As MyRF and Olig2 are the key transcription factors that orchestrate myelination (Bergles and Richardson, 2015; Yu et al., 2013), this suggests that the loss of endogenous ATM activity leaves the cell in an ambiguous state of differentiation. We propose that in response to the lowered levels of Olig2 in A-T, some terminally differentiated OLs begin to divide again. As a result, they enter a lethal cell cycle stall that leads to their death. As the reduction of Olig2 in OPC coincides with the astrogliosis, the combined effects of ATM on cell cycle and Olig2 may drive the substantial astrogliosis in A-T possibly through a transdifferentiation program (De Leon et al., 1976; Kanner et al., 2018; Sourander et al., 1966; Terplan and Krauss, 1969; Van de Kaa et al., 1994; Verhagen et al., 2012). These observations aligned with the molecular mechanism underlying the failure of *Atm*^-/-^ neural stem cells to differentiate into mOLs (Allen et al., 2001; Carlessi et al., 2009; Carlessi et al., 2013).

## Conclusion

The myelin abnormality in A-T are likely a cell autonomous effect of ATM deficiency in OL. The timing of the myelin/OL abnormalities and their quantitative relationship to different neuronal phenotypes are consistent with our hypothesis that the myelin defects are not simply a consequence of neuronal loss, but a direct loss of ATM function in the OL lineage. Our data demonstrate that the OL is highly vulnerable to loss of ATM activity at all stages of its life cycle.

## Version Notes

1. This version is based on reviewers’ recommendation to split the original manuscript (doi.org/10.1101/2021.01.22.20245217 deposited on January 26, 2021) into two medRxiv submissions. This first submission (deposited on 9^th^ November, 2023) focuses on the ATM deficiency in oligodendrocytes, and the second medRxiv submission (to be deposited in due course) will focuses on the effect of specific ATM mutation on oligodendrocyte differentiation.
2. We verify that this work is not under consideration for publication elsewhere, and that this publication is approved by all authors and all authors and tacitly or explicitly by the responsible authorities where the work was carried out, and that, if accepted, it will not be published elsewhere in the same form, in English or in any other language, including electronically without the written consent of the copyright holder.

## Authors’ contributions

K.H.T, A.C, performed animal experiments including histopathology, molecular tests, electron microscopy imaging with data acquisition and analysis with equal contribution. K.H.T, S.H.S.Y performed animal husbandry, oligodendrocyte cell culture and molecular tests. J.N.N. performed histopathology on postnatal tissues. G.W.Y.C performed animal husbandry. Q.W. performed cell line model experiment and immunocytochemistry B.Z. performed neuronal culture and analysis. Y.C and

L.J performed and facilitated electron microscopy. J.K., K.H.T and K.H. examined and analyzed the human tissues. K.H.T, A.C., S.H.S.Y and K.H. wrote the manuscript. K.H.T. and K.H. supervised the study and obtained funding, conceptualized the study, designed the experiment, and edited the final manuscript. All authors read, critically reviewed, and approved the manuscript.

## Conflict of Interests

All authors declare no conflict of interests in this manuscript.

## Data availability statement

The data that supports the findings of this study are available in the supplementary material of this article. Other data that support the findings of this study are available from the corresponding author upon reasonable request.

## Acknowledgments

The present work was generously supported by Health and Medical Research Fund (HMRF06173836 and HMRF04151436) of the Food and Health Bureau, Hong Kong Special Administrative Region, as well as National Institute of Neurological Disorders and Stroke (NINDS) R01NS120922 and the Pennsylvania Department of Health (4100087331). We acknowledge the kind support from the staff at NeuroBioBank of National Institutes of Health (NIH). All frozen human tissue in this study was obtained from the NIH NeuroBioBank at the University of Maryland, Baltimore, MD, and the PPFE tissues from Neuropathology Core, University of Pittsburgh Alzheimer’s Disease Research Center, United States. Dr Kofler and Alzheimer’s Disease Research Center at the University of Pittsburgh are supported by NIA P30 AG066468 and NIA P50 AG005133. Dr Jiang and the TEM work are supported by AoE/M-05/12.

---

**Fig. S1.**
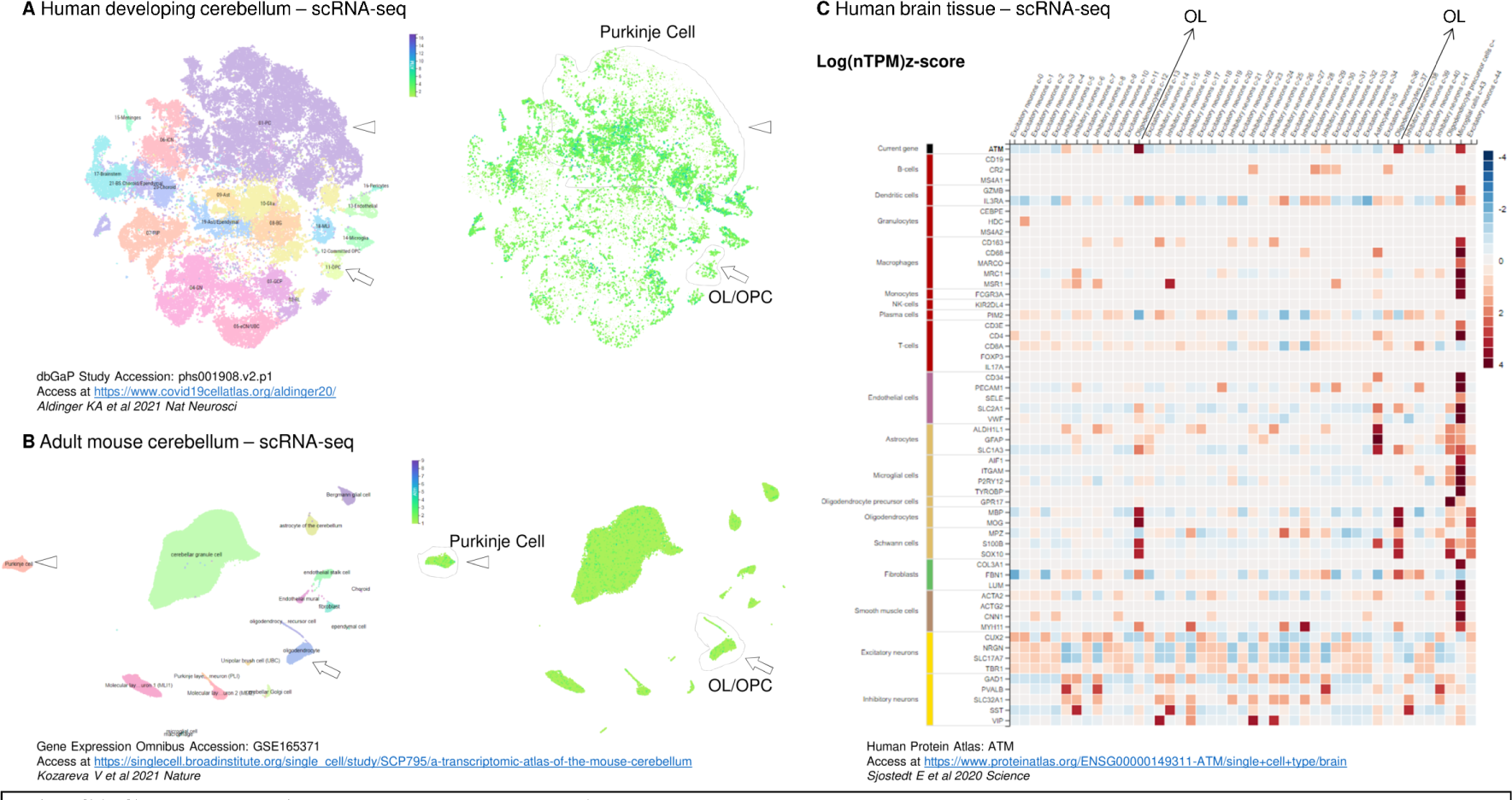
Cerebellar oligodendrocyte express ATM A, B. Single cell portal snapshots of tSNE map from two independent single cell RNA-seq (scRNA-seq) databases of cerebellum from developing human (Aldinger KA et al 2021 Nat Neurosci) and in adult mouse (Kozareva V et al 2021 Nature). The cell type-specific cluster distribution is annotated on *left*, with the *ATM* and *Atm* expression level denoted on the *right*. The expression level of Purkinje cells and OL/OPC population is circled. **C** Snapshot of scRNA-seq heatmap from Human Protein Atlas (Sjostedt E et al 2020 Science) of cell-type specific *ATM* expression level in different cell types (X axis, top row) in a brain tissue annotated against their gene set signature (Y axis). Of note, the expression level of ATM was the highest among microglia and OL population (arrow).

**Fig. S2.**
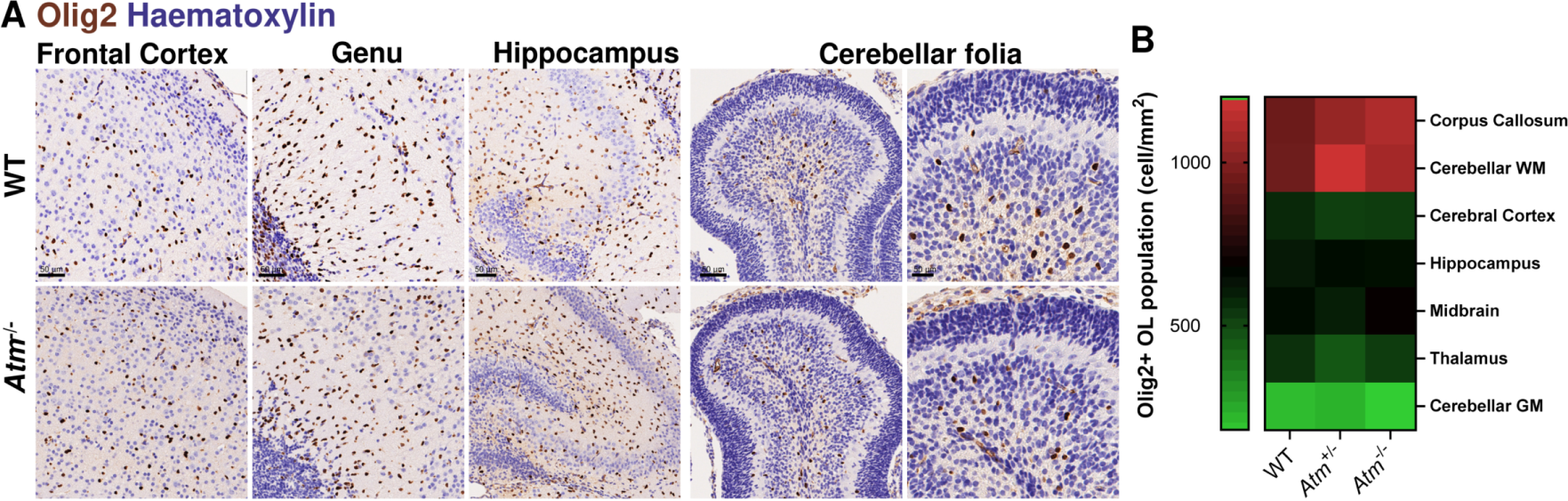
A. Representative immunohistochemistry image showing Olig2+ (DAB, brown) in the frontal cortex, genu of corpus callosum, hippocampus and cerebellum of WT and *Atm*-/- mouse brain at postnatal day 5. No discernible differences were found among regions between the two genotypes. **B** Quantification of showed a clear differences in Olig2 density between WM (corpus callosum and cerebellar WM) and GM (Cerebral cortex, hippocamps, midbrain thalamus and cerebellar GM, p < 0.0001), but no differences were detected among WT *Atm*+/- and *Atm*-/- (p = 0.759, Two-way ANOVA with HolmŠídák’s multiple comparisons test; n = 3 - 11).

**Fig. S3.**
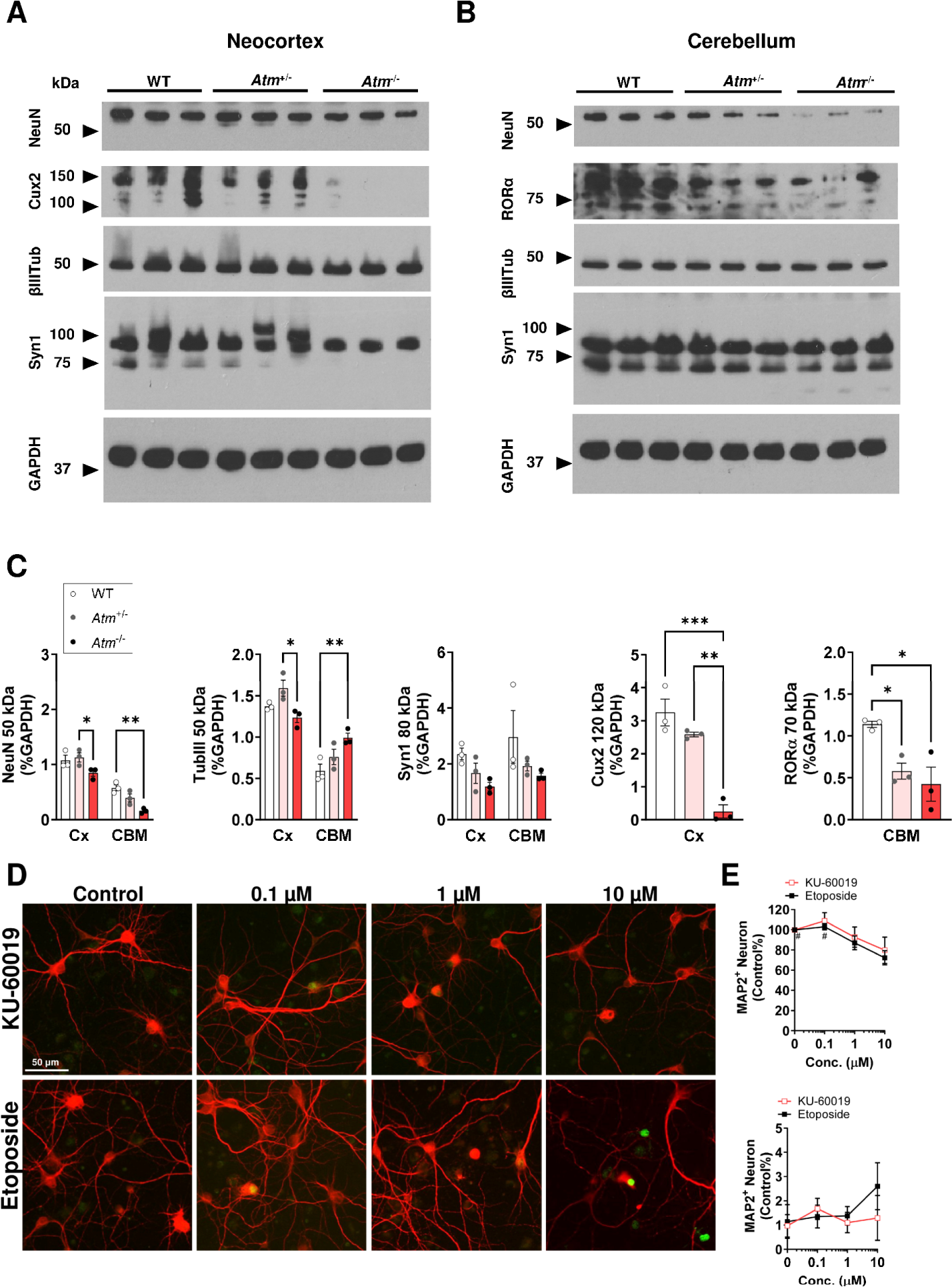
Immunoblots of neuron proteins (NeuN, βTubIII, Cux2 and RORα) and synaptic protein (Syn1) in the **A** neocortex and **B** cerebellum of WT *Atm*+/- and *Atm*-/- genotype at 1 month of age. **C** *Atm-*deficient genotype contributed to an overall significant changes of neuronal marker NeuN (p = 0.0011), cortical neuronal marker Cux2 (p = 0.0005), Purkinje cell marker RORα (p = 0.0195) and synaptic marker Syn1 (p = 0.034), but not axonal marker βTubIII, (p = 0.057) in neocortex an cerebellum. Two-way ANOVA with Holm-Šídák’s multiple comparisons test for two regions or one-way ANOVA with Tukey’s multiple comparisons test \**p* < 0.05, \*\**p* < 0.01, \*\*\**p* < 0.001 **D** Representative images of a dose response study of KU-60019-mediated and etoposide-mediated on cyclin D1 re-expression (green) in postmitotic neuronal culture (MAP2, red, WT embryonic) at 14 DIV. **E** Quantification showing the effect of ATM inhibition (red line) DSB-DNA damage (black line, 0 – 10 μM, 24 h), where etoposide, but not ATM inhibition, induced an overall reduction of the neuronal number in a dose-dependent fashion (p = 0.0046, one-way ANOVA, n = 4). etoposide also induced abnormal re-expression of cyclin D1 in neuron, but the increase did not reach statistical significance.

**Fig. S4.**
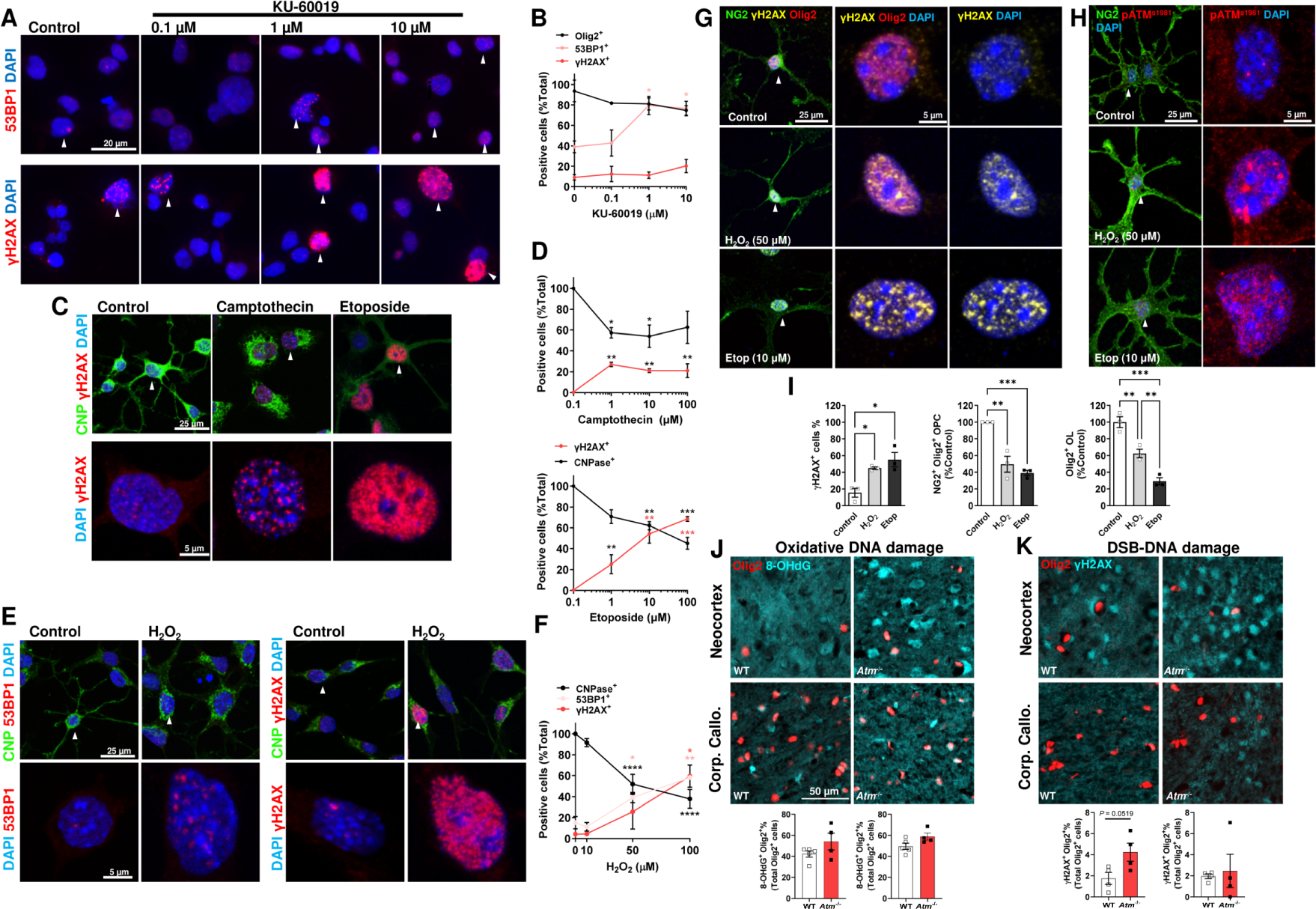
ATM inhibition, topoisomerase inhibition and oxidative stress induced DNA double strand breaks in OL A, B. Representative images and plot showing the inhibition of endogenous ATM activity by KU-60019 (0.1 - 10 μM, 24 h) increased the percentage of Oli-Neu cells with nuclear 53BP1+ foci (p = 0.015, red, top) and to a lesser extent γH2AX+ foci (red, bottom) without effect on Olig2+ cell number (p = 0.13, n = 4). **C, D** Representative confocal images and plots showing topoisomerase I and II inhibitors (Camptothecin and etoposide, respectively; 0.1 - 10 μM, 24 h) induced a significant increase of γH2AX+ foci (red) while reducing CNPase+ (green) cell number in Oli-Neu cells (Camptothecin, p = 0.0037; etoposide, p = 0.0003, n = 4). **E, F** Representative confocal images and plot showing H2O2 induced a significant increase of γH2AX+ (p = 0.012, not shown) and 53BP1+ foci (p = 0.0005, red) while reducing CNPase+ cell number (p < 0.0001) in Oli-Neu in a concentration dependent fashion. **G-I** In primary NG2+ OPC, H2O2-induced oxidative stress (50 μM, 24 h) and etoposide (10 μM, 24 h) also triggered a significant formation of γH2AX+ with increased expression of active ATM in the nucleus while reducing number of OPC. **J** Representative immunohistochemistry image of 8-OHG (8-hydroxyguanosine, cyan) and Olig2 (red) showed a high oxidative stress burden among OL population in WT and *Atm*-/- animals. **K** Representative immunohistochemistry image of γH2AX (cyan) and Olig2 (red) showed that such oxidative stress was associated with a trend of higher DSB formation of the OL in the cortex of the *Atm*-/- animals. One-way ANOVA with Tukey’s multiple comparisons test or unpaired t-tests \**p* < 0.05, \*\**p* < 0.01, \*\*\**p* < 0.001, \*\*\*\**p* < 0.0001. P values were colour coded on the does-response graphs.

## Notes

### Competing Interest Statement

The authors have declared no competing interest.

### Author Declarations

All experiments on post-mortem human brain tissues were approved by the Committee of Research Practices at The Hong Kong University of Science and Technology (HKUST) as well as Human Subjects Ethics Application Review board at The Hong Kong Polytechnic University (PolyU) All experimnets on animal models were approved by the Animal Ethics Committee, of the Committee on Research Practices of HKUST and Animal Subjects Ethics Sub-Committee of PolyU. All procedures were also conducted with a license from the Department of Health, Government of Hong Kong.

### Summary of Updates

This version is based on reviewers' recommendation to split the original manuscript (doi.org/10.1101/2021.01.22.20245217 deposited on January 26, 2021) into two medRxiv submissions. This first submission (deposited on 9th November, 2023) focuses on the ATM deficiency in oligodendrocytes, and the second medRxiv submission (to be deposited in due course) will focus on the effect of specific ATM mutation on oligodendrocyte differentiation.

