## Supplementary material for "Myelin pathology in ataxia-telangiectasia is the cell autonomous effect of ATM deficiency in oligodendrocytes": Table S1

| Antibody | Host & Clonality | Target | Application & dilution | Manufacturer | Cat No. (Clone) |
|---|---|---|---|---|---|
| 53BP1 | Rabbit Polyclonal | Marker of double strand breaks (DSB) damage in the nucleus | ICC: 1250 | Abcam | ab36823 (N/A) |
| 8-OHdG | Goat polyclonal | Marker of oxidative DNA lesion in the nucleus | IHC: 1:250 | EMD Millipore | AB5830 (N/A) |
| APC | Mouse monoclonal | Mature OL marker (commonly known as CC1) | IHC: 1:500 | Millipore | MABC200 (CC1) |
| ATM | Mouse monoclonal | Ataxia telangiectasia mutated DSB repair protein | IHC: 1:500<br>ICC: 1:250 | Abcam | ab78 (2C1) |
| Cleaved Caspase 3 | Mouse monoclonal | Apoptotic cells | IHC: 1:250 | EMD Millipore | MAB10753 (3D9.3) |
| Cux2 | Rabbit polyclonal | Marker for cortical neurons layer II-IIII | WB 1:1000 | Abcam | ab130395 |
| Cyclin A2 | Rabbit Monoclonal | Marker of G1/S transition | ICC: 1:250 | Abcam | ab181591 (EPR17351) |
| Cyclin D1 | Mouse monoclonal | A marker for G1/S phase transition during cell cycle | IHC: 1:250<br>ICC: 1:250 | Santa Cruz | sc-450 (72-13G) |
| Cyclin D1 | Rabbit polyclonal | A marker for G1 entry during cell cycle | IHC: 1:250 | Abcam | ab16663 (SP4) |
| GAPDH | Mouse monoclonal | Loading control for WB | WB: 1:10000 | Abcam | ab8245 (6C5) |
| GFAP | Rabbit polyclonal | Activated astrocytes | IHC: 1:500 | Abcam | ab7260 (N/A) |
| MAG | Mouse monoclonal | Myelin associated glycoprotein | WB: 1:1000 | Millipore | MAB1567 (N/A) |
| MAP2 | Chicken polyclonal | Neuronal/dendritic marker | IHC: 1:2000 | Abcam | ab5392 (N/A) |
| MAP2 | Mouse monoclonal | Neuronal/dendritic marker | WB: 1:5000 | Abcam | ab118853 (MT-07) |
| MBP | Mouse monoclonal | Myelin basic protein; Myelin; Myelinating OL | IHC: 1:1000<br>ICC: 1:500<br>WB: 1:2000 | Covance | SMI-99P (N/A) |
| MOG | Mouse monoclonal | Myelin Oligodendrocyte Glycoprotein | WB 1:1000<br>ICC 1:500 | Millipore | MAB5680 (N/A) |
| MyRF | Rabbit polyclonal | Perinuclear region and nucleus of postmitotic and myelinating OL | IHC: 1:250 | EMD Millipore | ABN45 (N/A) |
| NeuN | Mouse monoclonal | Pan-neuronal nuclei marker | WB 1:1000 | Millipore | MAB377 (A60) |
| NG2 (CSPG4) | Rabbit polyclonal | Marker of OL progenitor cells | WB: 1:500 | EMD Millipore | AB5320 (N/A) |
| Olig2 | Mouse monoclonal | Pan-OL-specific nuclear transcription factor | IHC: 1:250<br>WB: 1:500 | EMD Millipore | MABN50 (211F1.1) |
| pATM | Rabbit polyclonal | Ataxia telangiectasia mutated DSB repair protein phosphorylated at serine 1981 | ICC: 1:250 | Abcam | ab81292 (EP1890Y) |
| γH2A.X | Rabbit polyclonal | Marker of double strand breaks (DSB) Phosphorylated at serine 139 | IHC: 1:250<br>ICC: 1:250 | Abcam | ab2893 (N/A) |
| PLP | Rabbit polyclonal | Myelin proteolipid protein | WB 1:1000 | Thermofisher | PA3-150 (N/A) |
| pS/TQ | Rabbit polyclonal | Phospho-(Ser/Thr) ATM/ATR Substrate | WB 1:1000 | Cell Signaling | 2851S (N/A) |
| RORα | Rabbit polyclonal | Marker for purkinje cells in cerebellum | WB 1:1000 | Abcam | ab60134 (N/A) |
| Synapsin I | Rabbit polyclonal | Pre-synaptic marker | WB 1:1000 | Abcam | ab64581 (N/A) |
| Tubulin βIII | Mouse monoclonal | Neuron specific microtubule in axon | WB 1:1000 | BioLegend | MMS-435P (N/A) |
