## Supplementary material for "Myelin pathology in ataxia-telangiectasia is the cell autonomous effect of ATM deficiency in oligodendrocytes": Table S2

| Case | Age Range | Brain weight (g) | Sex | Group | PMI (hours) | *ATM* Variant (HGVSc) | ATM Variant (HGVSp) | InDels | Mutation type | ATM domain affected |
|---|---|---|---|---|---|---|---|---|---|---|
| AT1 | 25-30 | 1410 | F | Ataxia Telangiectasia | 4 | NM_000051.3:c.3712_3716delTTATT | NP_000042.3: p.Leu1238LysfsTer6 | deletion | frameshift_variant | N-terminal |
| AT2 | 30-35 | 1565 | M | Ataxia Telangiectasia | 13 | NM_000051.3:c.7875T>G | NP_000042.3: p.Asp2625Glu | snv | missense_variant | FAT/Kinase |
| AT3 | 15-20 | 1153 | F | Ataxia Telangiectasia | 2 | NM_000051.3:c.2284_2285delCT | NP_000042.3: p.Leu762ValfsTer2 | deletion | frameshift_variant | N-terminal |
| AT4 | 20-25 | 980 | F | Ataxia Telangiectasia | 2 | NM_000051.3:c.8880G>A | NP_000042.3: p.Trp2960Ter | snv | stop_gained | Kinase |
| AT5 | 15-20 | 1400 | M | Ataxia Telangiectasia | 15 | NM_000051.3:c.944delT | NP_000042.3: p.Leu315TyrfsTer5 | deletion | frameshift_variant | N-terminal |
| | | | | | | NM_000051.3:c.1564_1565delGA | NP_000042.3: p.Glu522IlefsTer43 | deletion | frameshift_variant | N-terminal |
| AT6 | 25-30 | 1380 | M | Ataxia Telangiectasia | 7 | NM_000051.3:c.3085dupA | NP_000042.3: p.Thr1029AsnfsTer19 | insertion | frameshift_variant | N-terminal |
| | | | | | | NM_000051.3:c.7327C>T | NP_000042.3: p.Arg2443Ter | snv | stop_gained | FAT |
| AT7 | 25-30 | N.D. | F | Ataxia Telangiectasia | 3 | NM_000051.3:c.6404_6405insTT | NP_000042.3: p.Arg2136Ter | insertion | frameshift_variant | FAT |
| AT8 | 20-25 | 1370 | M | Ataxia Telangiectasia | 15 | NM_000051.3:c.1290_1291delTG | NP_000042.3: p.Cys430Ter | deletion | frameshift_variant | N-terminal |
| AT9 | 25-30 | 1572 | M | Ataxia Telangiectasia | 14 | NM_000051.3:c.6404_6405insTT | NP_000042.3: p.Arg2136Ter | insertion | frameshift_variant | FAT |
| | | | | | | NM_000051.3: c.8036_8051 delATCTGGTGACTATACA | NP_000042.3: p.Asn2679SerfsTer9 | deletion | frameshift_variant | FAT/Kinase |
| AT10 | 35-40 | 1180 | M | Ataxia Telangiectasia | 20 | NM_000051.3:c.742C>T | NP_000042.3:p.Arg248Ter | snv | stop_gained | N-terminal |
| | | | | | | NM_000051.3:c.6095G>A | NP_000042.3:p.Arg2032Lys | snv | missense_variant, splice_region_variant | FAT |
| NC1 | 10-20 | 1600 | M | Control | 33 | | | | | |
| NC2 | 10-20 | 1660 | M | Control | 18 | | | | | |
| NC3 | 20-25 | N.D. | M | Control | 14 | | | | | |
| NC4 | 30-35 | 1550 | M | Control | 16 | | | | | |
| NC5 | 20-25 | 1160 | F | Control | 14 | | | | | |
| NC6 | 30-35 | 1800 | M | Control | 27 | | *Not applicable* | | | |
| NC7 | 15-20 | 1400 | F | Control | 5 | | | | | |
| NC8 | 25-30 | 1240 | F | Control | 11 | | | | | |
| NC9 | 25-30 | 1530 | M | Control | 12 | | | | | |
| NC10 | 15-20 | 1350 | F | Control | 16 | | | | | |
| NC-Age1 | 80-85 | N.D. | M | Ageing-Control | 5 | | | | | |
| NC-Age2 | 70-75 | N.D. | M | Ageing-Control | 3 | | | | | |
| NC-Age3 | 85-90 | N.D. | M | Ageing-Control | 7 | | | | | |
| NC-Age4 | 60-65 | N.D. | m | Ageing-Control | 6.5 | | | | | |
| NC-Age5 | 75-80 | N.D. | M | Ageing-Control | 6 | | *Not applicable* | | | |
| NC-Age6 | 75-80 | N.D. | F | Ageing-Control | 10.5 | | | | | |
| NC-Age7 | 70-75 | N.D. | F | Ageing-Control | 4.5 | | | | | |
| NC-Age8 | 85-90 | N.D. | M | Ageing-Control | 4 | | | | | |
| NC-Age9 | 75-80 | N.D. | M | Ageing-Control | 14 | | | | | |
